# Self-Supervised Pretraining Enables High-Performance Chest X-Ray Interpretation Across Clinical Distributions

**DOI:** 10.1101/2022.11.19.22282519

**Authors:** Niveditha S. Iyer, Aditya Gulati, Oishi Banerjee, Cécile Logé, Maha Farhat, Agustina D. Saenz, Pranav Rajpurkar

## Abstract

Chest X-rays (CXRs) are a rich source of information for physicians – essential for disease diagnosis and treatment selection. Recent deep learning models aim to alleviate strain on medical resources and improve patient care by automating the detection of diseases from CXRs. However, shortages of labeled CXRs can pose a serious challenge when training models. Currently, models are generally pretrained on ImageNet, but they often need to then be finetuned on hundreds of thousands of labeled CXRs to achieve high performance. Therefore, the current approach to model development is not viable on tasks with only a small amount of labeled data. An emerging method for reducing reliance on large amounts of labeled data is self-supervised learning (SSL), which uses unlabeled CXR datasets to automatically learn features that can be leveraged for downstream interpretation tasks. In this work, we investigated whether self-supervised pretraining methods could outperform traditional ImageNet pretraining for chest X-ray interpretation. We found that SSL-pretrained models outperformed ImageNet-pretrained models on thirteen different datasets representing high diversity in geographies, clinical settings, and prediction tasks. We thus show that SSL on unlabeled CXR data is a promising pretraining approach for a wide variety of CXR interpretation tasks, enabling a shift away from costly labeled datasets.

## Introduction

Chest X-rays (CXRs) are the most commonly used type of medical imaging globally, critical for the screening, diagnosis, and management of many life-threatening diseases. However, many medical systems currently lack the resources to accurately read CXRs, due to the time-consuming nature of CXR interpretation and an international shortage of radiologists^1,2^. Therefore, there have been significant efforts to create AI tools that can assist with CXR interpretation. Deep learning models have shown promise in this area, matching the performance of trained radiologists on a variety of tasks^3,4,5,6^.

In current practice, AI tools for any given CXR interpretation task are usually trained using a large CXR dataset with labels specifically addressing that task. Unfortunately, obtaining CXR data can itself be a costly, time-consuming process. Furthermore, obtaining high-quality labels requires experienced radiologists to manually interpret and annotate each CXR, which is extremely expensive^7^. The resulting lack of reliably labeled CXR data has posed a significant challenge for CXR model development, especially for less common tasks unaddressed by labels in existing datasets. For reference, one of the most frequently used datasets is CheXpert, a single-center dataset of 224,316 CXRs. While this dataset offers lower-quality “weak” labels automatically extracted from the radiology reports, board-certified radiologists manually annotated only 700 images.^8^ Our project aims to overcome the label shortage, addressing a key question: whether we train high-performing models without relying on large, densely annotated datasets.

We focus on transfer learning, a procedure that is widely used to improve model performance. In transfer learning, models are first pretrained on a large, general-purpose dataset and then finetuned on a more specific dataset addressing the target task. The pretraining step can compensate for weaknesses in downstream target datasets, such as when datasets are small or have weak labels. Over the past ten years, models for image interpretation have usually been pretrained specifically on ImageNet, a large dataset of natural images^9^; ImageNet pretraining has been successfully applied to a variety of medical image tasks, ranging from CXR interpretation to skin cancer classification to the diagnosis of retinal disease^10,11,12^. Unfortunately, the benefits of transfer learning lessen when the source data differs dramatically from the target data, as in the case of ImageNet images, which capture color images naturally occurring in the world, and CXRs, which are often taken in standard grayscale views with subtle local pixel differences accounting for differences in their classification.^13^ ImageNet-pretrained models often need to be finetuned on large, labeled CXR datasets containing tens or hundreds of thousands of datapoints in order to achieve high performance on downstream tasks. We hypothesize that pretraining on source datasets that more closely resemble target CXR datasets can improve downstream performance, potentially allowing CXR models to use significantly less labeled data during finetuning.

Self-supervised approaches, which train models without labels, have shown promising results on medical imaging tasks.^14^ For example, smaller scale studies have shown that self-supervised models pretrained on CheXpert had better performance and generalized better than models pretrained through supervised learning on ImageNet^15^. Furthermore, self-supervised models like MedAug have been shown to achieve high performance on specific chest X-ray pathology classification tasks using as little as 1% of CheXpert training dataset, making self-supervised learning (SSL) a promising approach to label-efficient training^16^. However, it is unknown whether self-supervised pretraining on unlabeled CXRs can outperform ImageNet-pretrained models across a range of CXR tasks.

To evaluate whether self-supervised models outperform ImageNet-pretrained models across clinical distributions, we implemented seven self-supervised learning approaches to pretrain models on a large CXR dataset without using labels. We compared the performance of these models against traditional Imagenet-pretrained models on thirteen datasets covering a variety of tasks, such as lung nodule detection, line and tube placement, edema severity classification, pediatric pneumonia detection, pneumothorax detection and multi-class differential diagnosis across demographics and clinical settings. After performing supervised learning on our target datasets, we evaluated how models behave across different distributions covering datasets of different sizes with different labeling methodologies.

Our results demonstrate that self-supervised pretrained models may learn generalizable chest X-ray representations, which improve performance over ImageNet-pretrained models; using such models can enable a shift away from costly labeled datasets for a large variety of chest-X-ray interpretation tasks across different clinical distributions. By eliminating the need to train models on large labeled datasets that are difficult to obtain, this work will enable institutions to more rapidly train and evaluate models across distinct clinical tasks, getting a step closer to model deployment into clinical settings.

## Results

### Experimental Setup

To evaluate whether self-supervised CXR pretraining outperforms ImageNet pretraining, we selected seven SSL methods, three that only use CXR images (MedAug^16^, S2MTS2^17^, MoCo-CXR^18^) and four that use both chest X-ray images and corresponding radiology reports (CXR-RePaiR-CLIP^19^, ConVIRT^20^,REFERS^21^, GLoRIA^22^). We give further details on these learning algorithms in the Methods section. Significantly, none of these methods used any labels, requiring only CXRs, reports, and metadata that were naturally produced as part of medical workflows.

To assess whether models can generalize across clinical distributions, we chose a wide variety of downstream CXR datasets that we used to finetune and validate our models. These datasets came from diverse sites, including Brazil^23^, China^24,25^, United States^,25,,26,27,28,29,30,31^, Spain^32^, Japan^33^, Vietnam^34^ and other countries in Eastern Europe and Central Asia.^35^ They also addressed a large set of tasks, such as lung nodule detection, line and tube placement, edema severity classification, pediatric pneumonia detection, pneumothorax detection and multi-class differential diagnosis; some datasets address wide-ranging detection tasks for multiple pathologies. Furthermore, these datasets were collected in a variety of clinical settings, including a pediatric setting (Pediatric Pneumonia); a Tuberculosis screening program (Montgomery); an emergency department (MIMIC CXR); outpatient (Shenzhen) and in-hospital settings (see Table 1).

**Table 1:**
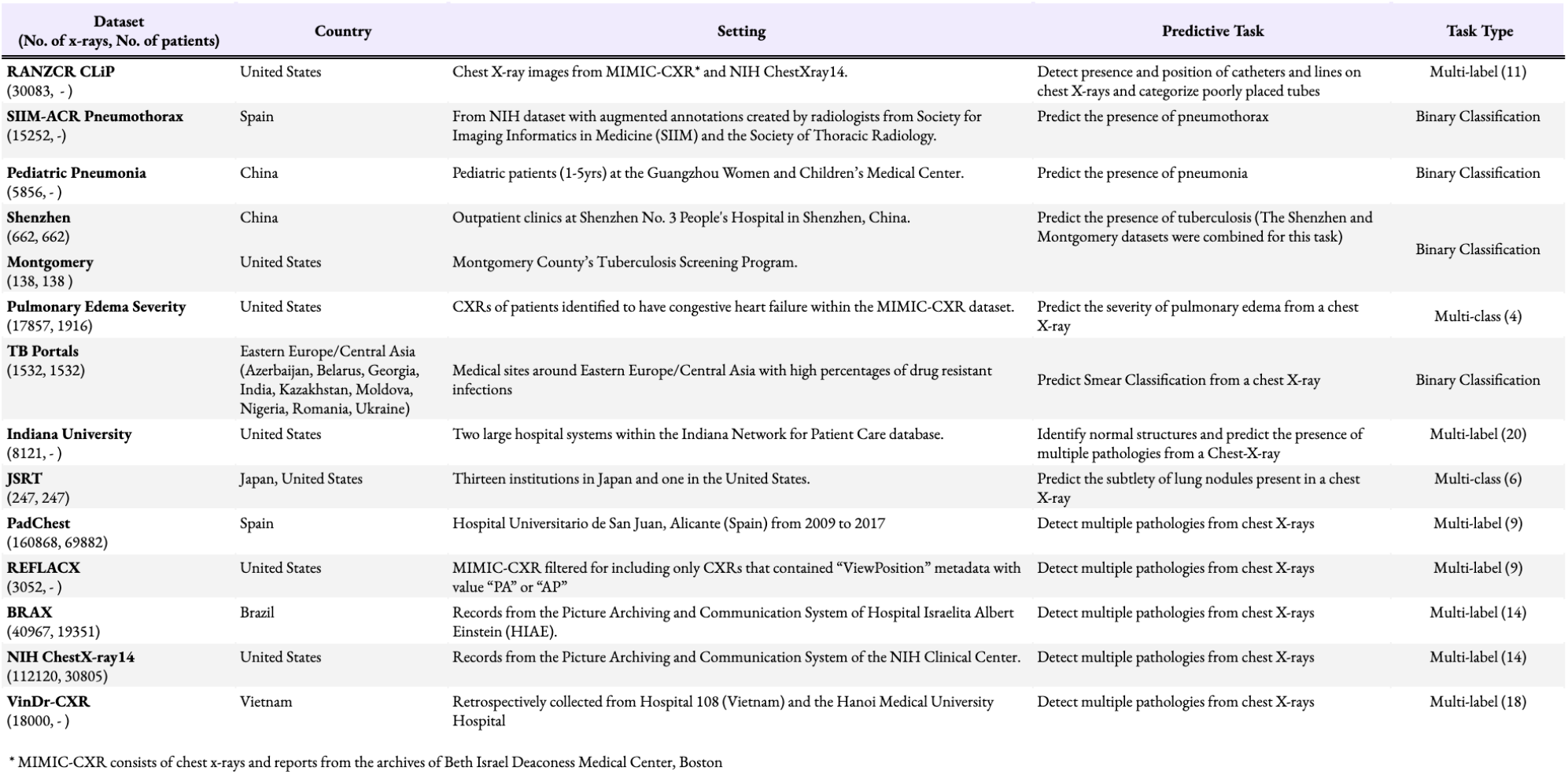
Overview of each target dataset: number of radiographs, number of unique patients and description of the patient population (country and setting), descriptions of prediction task and task type for each dataset. We selected thirteen distinct datasets that vary in size, patient population (age, gender, country) and the type of predictive task.

Once pretrained, models were adapted for each downstream task, using all available training data in the target dataset. In our first set of experiments, we fine tuned all model parameters and compared the performance of SSL CXR-pretrained models against the ImageNet-pretrained models, controlling for the model capacity and hyperparameter search (as detailed in Methods). In the second set of experiments, we performed linear probing, freezing the weights of our CXR pretrained models and training one final fully-connected classifier layer for each. These experiments directly evaluated the quality of the features learned through CXR self-supervision. After training, we tested the performance of our models using the testing data from each dataset.

### Self-supervised CXR pretrained models outperform ImageNet pretrained models across clinical distributions

We found that finetuned SSL models consistently provided improvements over ImageNet pretraining across datasets (see Table 2). The complete tables showing these results can be found in Supplementary Note 3. In particular, the highest improvements in the average AUC achieved across tasks were seen on the NIH dataset with ConVIRT (0.108 improvement; [95 % CI 0.082, 0.1333]) and on the REFLACX dataset, also with ConVIRT (0.103 improvement, [95 % CI [0.035,0.181]). On both datasets, the highest AUC for each task (each pathology/condition in a dataset) was obtained by a self-supervised model; on the NIH dataset, the highest improvement was seen when identifying atelectasis (0.250 improvement [95% CI 0.224,0.278] with ConVIRT), while on the REFLACX dataset, the highest improvement was seen for pneumothorax detection (AUC improvement of 0.256 [95% CI 0.118, 0.437] with GLoRIA).

**Table 2:** Maximum AUC improvements over ImageNet pretraining achieved across the seven self-supervised models for each dataset. Starred entries indicate statistically significant improvements (p < 0.05) over the ImageNet Pretrained baseline. SSL models outperformed ImageNet pretrained models across datasets with distinct clinical tasks and patient populations.

| Dataset | ImageNet-Pretrained Baseline | Maximum Improvement over ImageNet AUC with Linear Probing and Finetuning |  |
| --- | --- | --- | --- |
|  |  | Linear Probing | Finetuning |
| RANZCR CLiP | 0.813 [0.791, 0.833] | 0.024 [0.002, 0.049]* | 0.071 [0.045, 0.097]* |
| SIIM-ACR | 0.917 [0.905, 0.929] | 0.001 [-0.011, 0.016] | 0.017 [0.006, 0.029]* |
| Pediatric Pneumonia | 0.930 [0.898, 0.953] | 0.064 [0.040, 0.094]* | 0.053 [0.029, 0.081]* |
| Shenzhen & Montgomery | 0.923 [0.876, 0.967] | 0.033 [-0.014, 0.076] | 0.046 [-0.001, 0.099] |
| MIMIC CXR | 0.713 [0.685, 0.740] | 0.078 [0.058, 0.098]* | 0.089 [0.060, 0.119]* |
| TB Portals | 0.707 [0.665, 0.766] | 0.008 [-0.069, 0.074] | 0.008 [-0.060, 0.070] |
| Indiana | 0.727 [0.650, 0.798] | 0.095 [0.009, 0.176]* | 0.072 [-0.001, 0.150] |
| JSRT | 0.543 [0.347, 0.709] | 0.044 [-0.175, 0.257] | 0.015 [-0.182, 0.204] |
| PadChest | 0.635 [0.573, 0.694] | -0.052 [-0.137, 0.035] | 0.024 [-0.041, 0.096] |
| REFLACX | 0.744 [0.674, 0.804] | 0.109 [0.037, 0.190]* | 0.103 [0.035, 0.181]* |
| BRAX | 0.779 [0.734, 0.821] | 0.042 [0.000, 0.087] | 0.041 [0.003, 0.077]* |
| NIH ChestX-ray14 | 0.706 [0.683, 0.731] | 0.091 [0.061, 0.118]* | 0.108 [0.082, 0.133]* |
| VinDR-CXR | 0.814 [0.780, 0.847] | 0.064 [0.027, 0.102]* | 0.065 [0.028, 0.103]* |

Figure 1 shows a per-task comparison across all multi-label datasets between each dataset’s best SSL model and the ImageNet-pretrained model.

**Figure 1.**
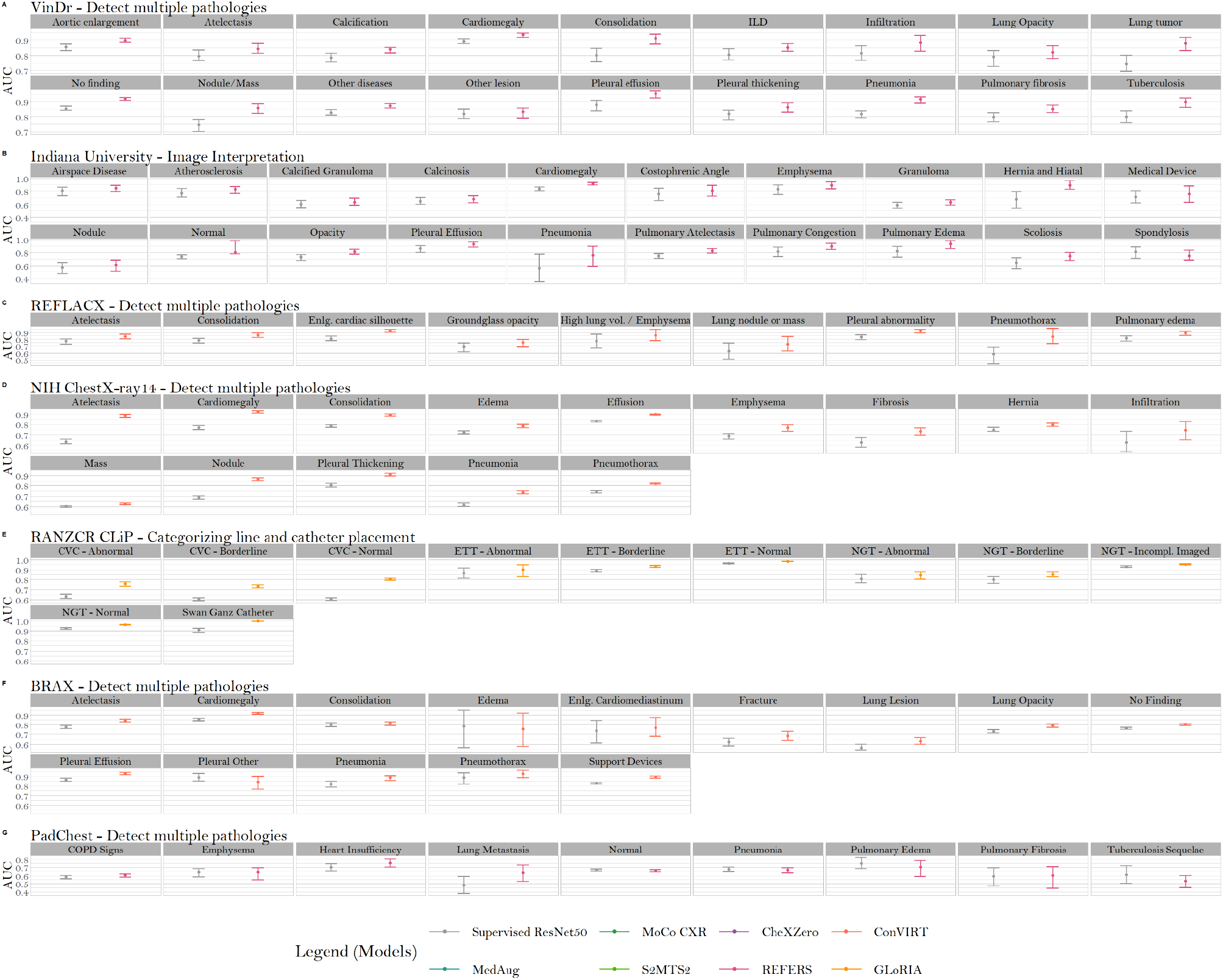
AUC score breakdown by task for the seven multi-label datasets with finetuning. On each task, we compare the AUC of the dataset’s overall best-performing model against that of the ImageNet-pretrained model. Across datasets, SSL models had a great performance detecting cardiomegaly, pleural effusion, pneumothorax and ETT placement.

We found that with linear probing, SSL models outperformed the finetuned ImageNet model on twelve of thirteen datasets (see Table 2). The largest average improvements in performance across tasks were seen on the REFLACX dataset with GLoRIA (0.109 difference over the supervised model, [95% CI 0.037, 0.190]) and on the NIH ChestX-ray14 dataset with REFERS (0.091 difference, [95% CI 0.061,0.118]). On both datasets, the highest AUC per task was achieved by a self supervised CXR pretrained model. On the pathologies within REFLACX, the largest performance improvement was achieved by GLoRIA when recognizing pneumothorax (0.215 improvement, [95% CI 0.068,0.412] over the ImageNet-pretrained model after finetuning). Within the NIH dataset, the largest improvement was seen with CXR-RePaiR-CLIP when identifying atelectasis (0.244 improvement, [95% CI 0.221,0.266]). PadChest is the only dataset where the finetuned ImageNet pretrained model outperformed all self-supervised models after linear probing, though its improvement was not statistically significant: the AUC of the best-performing self-supervised model (CXR-RePaiR-CLIP) was lower by only -0.005 [95 % CI-0.088,0.073].

### Comparing Different SSL Methods

To evaluate whether pretraining on images and text together yielded an advantage over image-only methods, we compared the average performance across all datasets of image-text with image-only SSL models with finetuning and linear probing. To further assess whether finetuning significantly boosted SSL models’ performance, for both image-text and image-only methods, we compared each model’s average performance across all datasets between finetuning and linear probing.

We found that finetuned SSL models that used both images and text outperformed the image-only self-supervised methods (See Figure 2 and Table 4 of Supplementary Note 3). On four of the thirteen datasets, the highest average AUC was achieved by GLoRIA and ConVIRT. The best-performing (highest average finetuning improvement across all datasets) image-text methods were ConVIRT (AUC of 0.810) and REFERS (AUC of 0.810), closely followed by GLoRIA (AUC of 0.799). The best-performing image-only method was S2MTS2 (AUC of 0.781).

**Figure 2.**
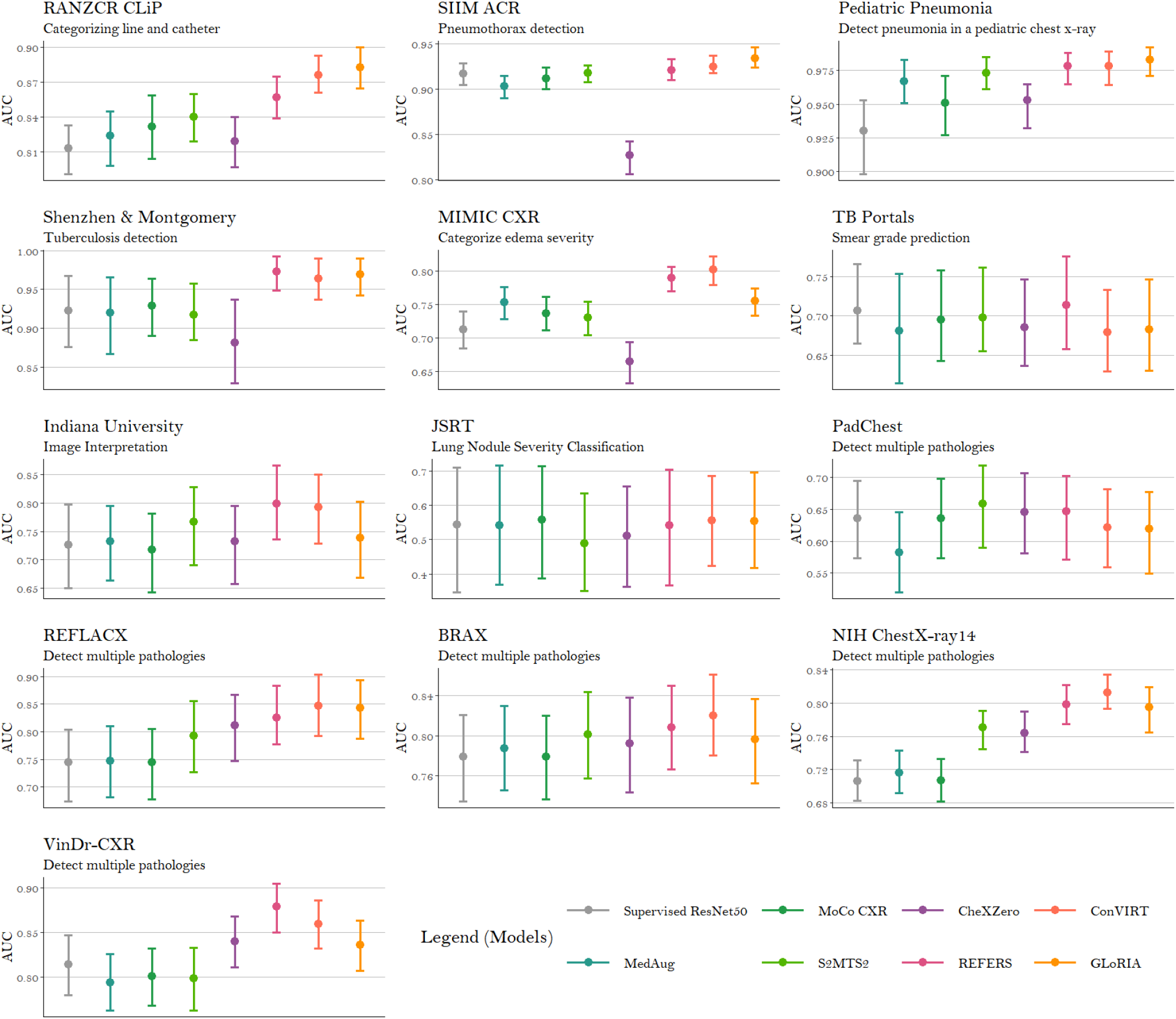
**Finetuning** AUC (averaged across tasks for datasets containing multiple tasks) achieved by finetuning of each ImageNet-pretrained model and seven self-supervised models on thirteen datasets. The figure shows the AUC of each SSL model and the ImageNet-pretrained model. SSL models that used both image and text outperformed image only SSL models.

We found that, with linear probing, all image-text SSL models outperformed image-only SSL models averaged across datasets. (See Figure 3 and Table 5 of Supplementary Note 3). REFERS had the best average performance (AUC 0.802), followed closely by ConVIRT (average AUC 0.792), CXR-RePaiR-CLIP (average AUC 0.789) and GloRIA (average AUC 0.783). All three image-only SSL models underperformed the finetuned ImageNet-pretrained model. The best image-only SSL model was MedAug, which achieved an average AUC of 0.724 across datasets.

**Figure 3.**
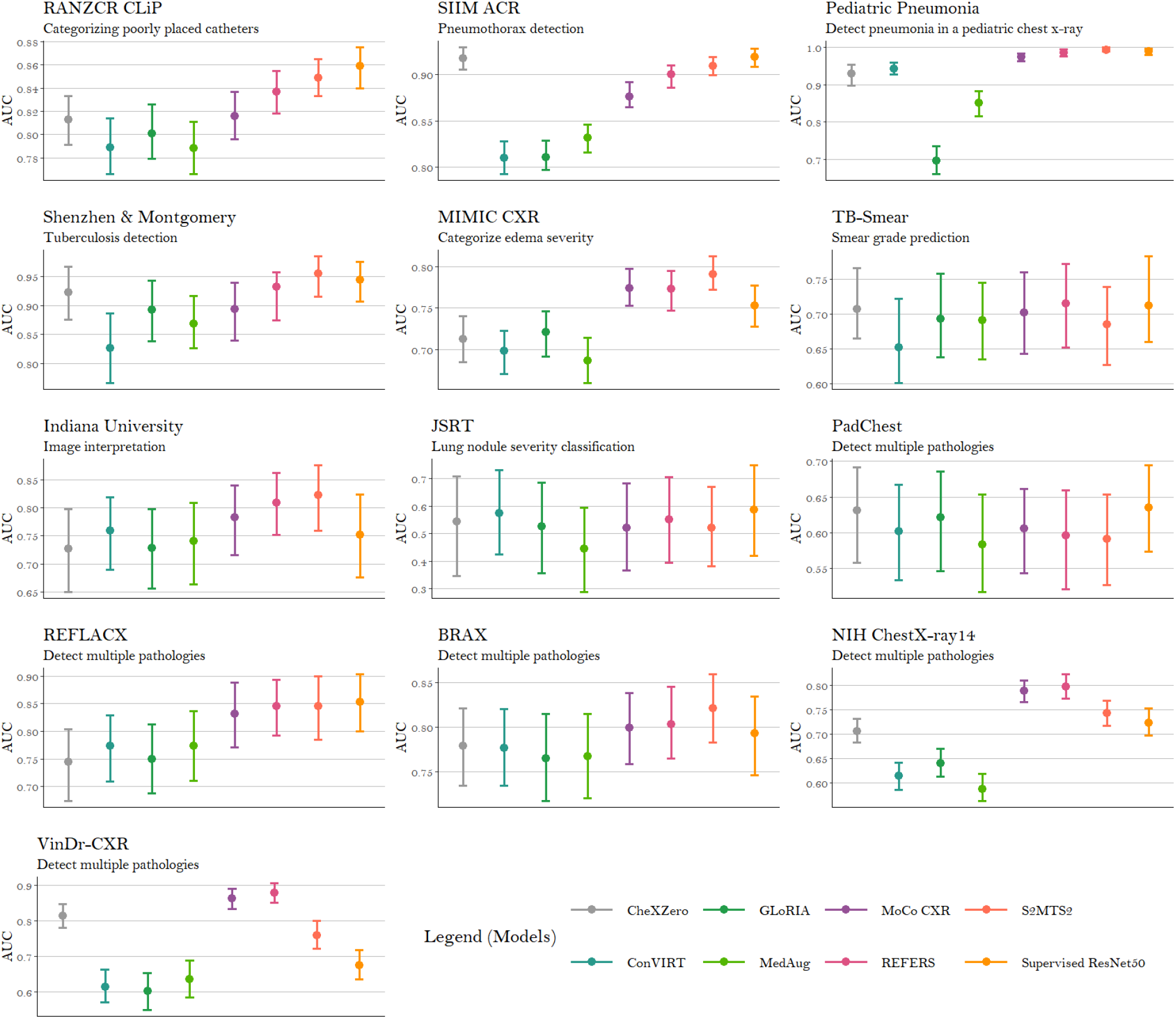
**Linear Probing** AUC (averaged across tasks for datasets containing multiple tasks) achieved through linear probing of each ImageNet-pretrained model and seven self-supervised models on thirteen datasets. (A) to (M) figure shows the AUC of each SSL model and the ImageNet-pretrained model. SSL models that used both image and text outperformed image only SSL models.

We found that the best linear probing models outperformed the best finetuned models on six out of thirteen datasets. However, model performance on average across datasets was better after finetuning. Only CXR-RePaiR-CLIP had a higher performance with linear probing (0.025 average improvement over finetuning). The three models that benefited the most from finetuning were pretrained only on images: S2MTS2 (0.069 difference over linear probing), MoCo-CXR (0.059), and MedAug (0.041).

## Discussion

### Performance across distribution shifts

Several studies have demonstrated a significant drop in performance across a variety of CXR applications when deep learning systems were tested on datasets outside their training domain^36,37,38^. Machine learning models trained through supervised learning often achieve high performance on only a single dataset due to multiple factors, such as technical differences in how CXR images are captured across datasets and varying disease distributions across different populations^37^. We demonstrated that the best self-supervised models maintained their performance across datasets from different geographies, populations and tasks. Although there was not a single model that maintained high performance across all thirteen datasets, our results are promising, suggesting that self-supervised pretraining can broadly improve generalizability. We believe that labeled data is still essential for high performance and generalization, since both supervised and self-supervised models had consistently poor performance across PadChest and JSRT both for finetuning and linear probing.

### Self-supervised models learn better representations using both images and reports

Image-text models learned higher-quality, more generalizable representations than image-only models, across both our finetuning and linear probing experiments. Amongst the image-text models, ConVIRT and REFERS were generally the best-performing models after both finetuning and linear probing. However, they did not achieve the highest performance on all datasets, indicating that while certain models can learn more useful representations on average, they may still falter on specific tasks. Our results did not find any one SSL model that consistently outperforms other models on every new dataset / new task, and work remains to be done on this front.

### Clinical Implications

Although the performance of CXR self-supervised models was overall better than ImageNet-pretrained supervised models, results were not homogeneous across different tasks and datasets.

For tasks that assess the placement of lines and tubes, as seen on the the RANCZR CLiP dataset, CXR-pretrained models generally did better than the ImageNet-pretrained model. Both kinds of models could successfully detect NGT or ETT placement, but neither model did well enough to evaluate central line placements. The latter is a more complicated task, because this dataset includes different types of central catheters (central line, peripherally inserted central catheter, Ports, Hickman’s) and because these catheters can be inserted at different levels and from either the right or left side of the body. Performance likely dropped due to the increased complexity of the task.

For complex tasks with multiple pathologies, we observed that our models performed poorly on datasets with multiple labels that did not clearly distinguish between a specific finding and a diagnostic impression. Specifically, they underperformed on PadChest when distinguishing between heart insufficiency and pulmonary edema or emphysema and chronic obstructive pulmonary disease (COPD), even though PadChest is a relatively large dataset with more than 160,000 images. These labels can be particularly difficult to apply, and even the most experienced radiologists disagree on the correct label due to ambiguity in instructions. These findings contribute to the hypothesis that size is not the only important feature of a dataset; it is also necessary to have clear, specific labels, especially for complex tasks.

When the task involves a more subjective appraisal, as when determining the severity of pulmonary edema or of subtlety of a pulmonary nodule, we found that neither set of models did well. These results mirror clinical practice, where board-certified radiologists and emergency medicine physicians demonstrate a sensitivity of 77% and 59% respectively, when detecting the presence of edema, and their inter-agreement is also low^39,40^. Similarly, when detecting lower-subtlety pulmonary nodules, performance decreases even among experienced radiologists^41^.

Although more research is necessary, self-supervised pretraining is a promising approach to image interpretation tasks, especially for tasks where datasets are small but contain clear, specific labels. A recent study found that pretraining the data on natural images, followed by pretraining on large weakly labeled CXR datasets, and finally, task-specific training on small labeled datasets could significantly reduce label requirements on the target dataset^42^; thus the composition of large-scale out-of-domain pretraining and self-supervised pre training may be a fruitful direction for future work.

Overall, CXR self-supervised pretrained models did better than ImageNet-pretrained models, especially for image-text models. There was no task where the ImageNet-pretrained model consistently did better than models pretrained through CXR self-supervision. Tasks where CXR self-supervised models did particularly well included the detection of pneumothorax, cardiomegaly, pleural effusion, and tube placement. Within these tasks, models maintained high performance across different populations and clinical settings, indicating that these models could be ready for deployment, assisting clinicians in identifying those critical conditions instantly. It is also important to note that the quality of the labels may influence performance more than the number of images and labels in a specific dataset; this is an important consideration when creating future datasets.

### Conclusion

In this study, we tested whether the emerging paradigm of self-supervised pretraining would outperform the traditional paradigm of ImageNet pretraining on different chest x-ray interpretation tasks. Traditionally, ImageNet pretraining has been the dominant approach to tackling medical imaging tasks with limited data. However, our results indicate that self-supervised pretraining on medical data could take over as the traditional paradigm for low-data medical tasks. In addition to significantly improving performance across downstream tasks, this study represents a step forward in learning generalizable representations of chest X-rays. Such self-supervised pretrained models could be easily adapted to and deployed in diverse clinical settings.

## Methods

### Model Descriptions

We benchmarked the performance of seven SSL methods. The methods were primarily divided into two categories, depending on whether the pretraining tasks used images alone or used both images and reports. The image-only models (MedAug, S2MTS2, MoCo-CXR) were pretrained on CXR images, whereas image+reports models (CXR-RePaiR-CLIP, ConVIRT, REFERS, GLoRIA) were pretrained on the CXR images along with corresponding reports. These models used different backbone architectures (ResNet-50^43^, CLIP-ViT^44^, Dense121^45^) and underlying pretraining frameworks (contrastive learning^46^, CLiP, transformers^47^, self-attention^48^, student-teacher^49^, cross-supervision^50^). A detailed description of these methods is given in Table 3.

**Table 3.**
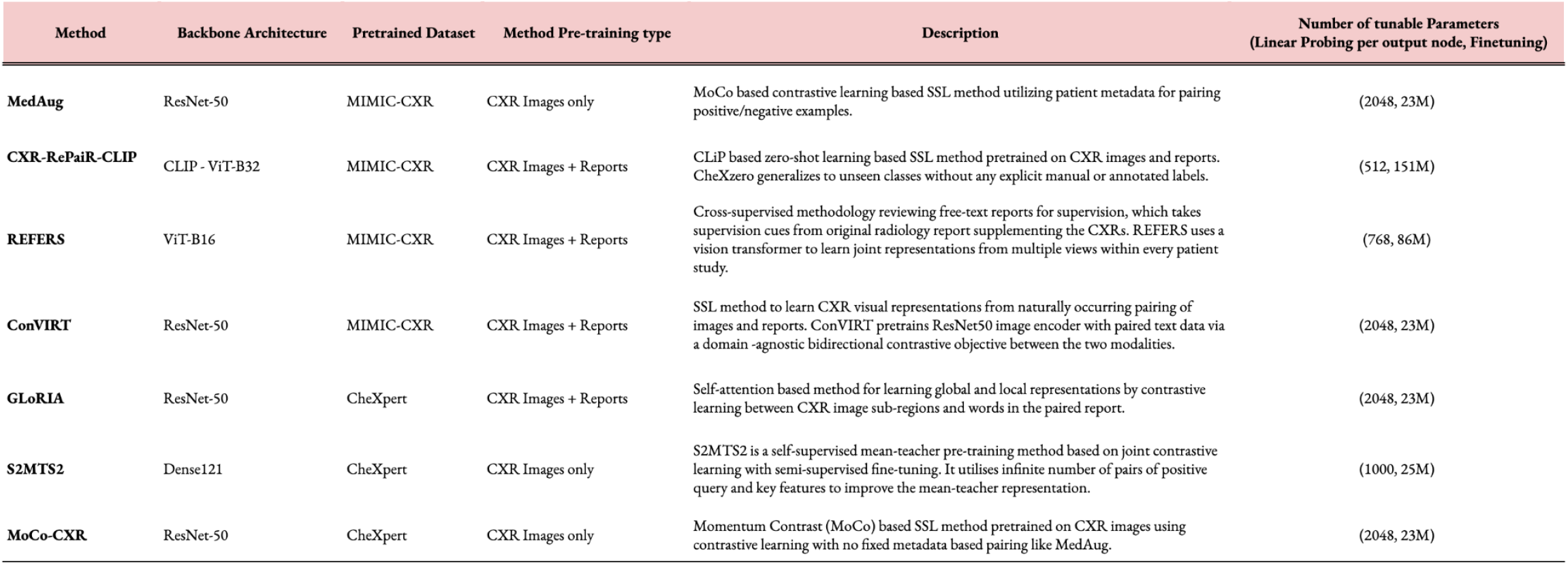
Overview of SSL methods with backbone architecture, pretraining dataset, pretraining type, description and number of tunable parameters during linear probing and finetuning.

| Method | Backbone Architecture | Pretrained Dataset | Method Pre-training type | Description | Number of tunable Parameters<br>(Linear Probing per output node, Finetuning) |
| --- | --- | --- | --- | --- | --- |
| **MedAug** | ResNet-50 | MIMIC-CXR | CXR Images only | MoCo based contrastive learning based SSL method utilizing patient metadata for pairing positive/negative examples. | (2048, 23M) |
| **CXR-RePaiR-CLIP** | CLIP - ViT-B32 | MIMIC-CXR | CXR Images + Reports | CLiP based zero-shot learning based SSL method pretrained on CXR images and reports. CheXzero generalizes to unseen classes without any explicit manual or annotated labels. | (512, 151M) |
| **REFERS** | ViT-B16 | MIMIC-CXR | CXR Images + Reports | Cross-supervised methodology reviewing free-text reports for supervision, which takes supervision cues from original radiology report supplementing the CXRs. REFERS uses a vision transformer to learn joint representations from multiple views within every patient study. | (768, 86M) |
| **ConVIRT** | ResNet-50 | MIMIC-CXR | CXR Images + Reports | SSL method to learn CXR visual representations from naturally occurring pairing of images and reports. ConVIRT pretrains ResNet50 image encoder with paired text data via a domain -agnostic bidirectional contrastive objective between the two modalities. | (2048, 23M) |
| **GLoRIA** | ResNet-50 | CheXpert | CXR Images + Reports | Self-attention based method for learning global and local representations by contrastive learning between CXR image sub-regions and words in the paired report. | (2048, 23M) |
| **S2MTS2** | Dense121 | CheXpert | CXR Images only | S2MTS2 is a self-supervised mean-teacher pre-training method based on joint contrastive learning with semi-supervised fine-tuning. It utilises infinite number of pairs of positive query and key features to improve the mean-teacher representation. | (1000, 25M) |
| **MoCo-CXR** | ResNet-50 | CheXpert | CXR Images only | Momentum Contrast (MoCo) based SSL method pretrained on CXR images using contrastive learning with no fixed metadata based pairing like MedAug. | (2048, 23M) |

### Adapting Strategies

We adapted self-supervised models in two ways: linear probing and finetuning. For linear probing, we added a logistic regression layer on top of the last layer of the backbones for 100 epochs, using the L2 norm and Limited-memory Broyden–Fletcher–Goldfarb–Shanno solver. For finetuning, we tuned all the model parameters with SGD optimizer for 100 epochs with a learning rate of 0.001, a batch size of 32, and a learning rate scheduling step of 200 with cosine annealing. These sets of hyperparameters are given in the official implementation of respective methods. Model parameter selection was based on the model with the highest AUC on the validation set. We ran the finetuning of SSL methods on a single NVIDIA RTX A4000 GPU card.

### Dataset Selection

We evaluated CXR self-supervised models across a range of tasks, using fourteen distinct datasets that vary in size, patient population, geography, setting, and type of predictive task. These tasks are a mix of binary classification (5 datasets), multi-label (7 datasets) and multi-class (2 datasets) tasks. The tasks that we have selected are described in Table 1.

### Dataset Processing

We split every dataset except the Pediatric Pneumonia and VinDr-CXR datasets into training, validation and test sets with 60%, 20%, and 20% of datapoints respectively. We ensured that there was no patient overlap between the three sets. The Pediatric Pneumonia and VinDr-CXR datasets were split into (80%, 10%, 10%) and (66%, 16%, 16%) respectively, as we retained the test split provided by the creators of the dataset.

### Statistical Analysis

We evaluated the performance of models using AUC scores, representing the area under the receiver operating characteristic curve. We reported the variability in our estimate of this measure using the percentile bootstrap: we constructed bootstrap confidence intervals by taking 1000 replicates of sampling test data points with replacement.

The nonparametric bootstrap was used to estimate the variability around each of the performance measures; 1000 bootstrap replicates from the test set were drawn, and each performance measure was calculated for the ImageNet-pretrained baseline model and the seven self-supervised models on these same 1000 bootstrap replicates. This process produced a distribution for each estimate, and the 95% bootstrap percentile intervals (2.5th and 97.5th percentiles) are reported. To find the performance improvement provided by the best SSL model over the ImageNet supervised learning model, we computed the differences of 1000 replicated AUCs and reported the 95% bootstrap percentile intervals^51^. The analyses were performed using statsmodels, scikit-learn and SciPy packages in Python.

### Package description - *cxrlearn*

The SSL methods implemented in this work were developed by multiple research groups across different platforms with multiple dependencies. We have integrated all SSL codebases into one reusable library called *cxrlearn. cxrlearn* provides documented functions for dataset conversion, self-supervised learning, model object generation, finetuning, linear probing, and evaluation. Our open-source library codebase is available here.

## Data Availability

Most datasets used in this study are public and can be accessed through their respective websites. Requests concerning the TB portals dataset should be addressed to Maha Farhat.

https://www.nature.com/articles/s41597-022-01608-8

https://www.sciencedirect.com/science/article/pii/S0092867418301545

https://www.ncbi.nlm.nih.gov/pmc/articles/PMC4256233/

https://www.nature.com/articles/s41597-021-01066-8

https://www.kaggle.com/jesperdramsch/siimacrpneumothorax-segmentation-data.

https://arxiv.org/abs/2109.14187

https://openaccess.thecvf.com/content_cvpr_2017/html/Wang_ChestX-ray8_Hospital-Scale_Chest_CVPR_2017_paper.html

https://academic.oup.com/jamia/article/23/2/304/2572395

https://www.nature.com/articles/s41597-019-0322-0

https://bimcv.cipf.es/bimcv-projects/padchest/

## Acknowledgements

PR was supported by a Harvard Data Science Initiative Competitive Research Fund Grant and by the NHLBI BioData Catalyst Fellowship.

## Contributions

Concept and design: PR, NI, AG. Acquisition, analysis, or interpretation of data: AG, NI, PR, AS, CL, MF. Drafting of the manuscript: NI, AS, AG, OB. Critical revision of the manuscript for important intellectual content: All authors. Statistical analysis: AG, NI. Obtained funding: PR. Administrative, technical, or material support: PR. Supervision: PR. All authors approved the final version.

## Corresponding author

Correspondence to Pranav Rajpurkar, PhD.

## Ethics declarations

The authors declare no competing interests.

