## Supplemental Material for "Self-Supervised Pretraining Enables High-Performance Chest X-Ray Interpretation Across Clinical Distributions"

### Extended data

#### Supplementary Note 1

Dataset descriptions

1. RANCZR CLiP Catheter and Line Positions

This dataset contains 30,086 chest X-rays selected from the NIH ChestX-ray14 dataset using search terms related to ETT, NGT, and CVC catheters. The annotations were produced by 43 labelers, including radiologists, radiology trainees and hospital medical officers. The data categorizes  the placement of Endotracheal Tubes (ETT - abnormal, borderline, normal), Nasogastric Tubes (NGT - abnormal, borderline, incompletely imaged, normal), Central Venous Catheters (CVC - abnormal, borderline, normal) and Swan Ganz Catheters (present, absent). Each of the categories is considered a label, giving us a total of 11 labels which were predicted upon. Note that a single image can have multiple labels (e.g. both ETT-Normal and ETT-Abnormal). We perform 11 binary classification tasks on this dataset, predicting whether each of the 11 labels applies to a given CXR.

2. SIIM Pneumothorax

This dataset is from the SIIM-ACR Pneumothorax Segmentation Competition on Kaggle. The images in the dataset come from the public NIH chest radiograph dataset with additional annotations. The images are stored in DICOM format, and annotations consist of run-length-encoded (RLE) masks, which indicate the location of the pneumothorax. The dataset annotations were provided by members of the SIIM Machine Learning Committee. We use a total of 12,089 unique X-rays and 12,748 RLEs. In the dataset, all images without pneumothorax are assigned an RLE mask value of -1. We assign the label “0” to all such images. The remaining images with RLE masks (and hence with pneumothorax) are assigned the label “1”. We performed binary classification on this dataset, predicting whether pneumothorax is present in each CXR.

3. Pediatric Pneumonia Chest X-ray

This dataset consists of 5856 CXRs of pediatric patients compiled from retrospective cohorts of pediatric patients between the ages of one and five years old from Guangzhou Women and Children’s Medical Center in Guangzhou, China. The dataset contains 4273 CXRs images characterized as showing pneumonia (2780 bacterial and 1493 viral) and 1583 normal CXRs. We performed binary classification on this dataset, predicting whether pneumonia is present in each CXR.

4. Shenzhen and Montgomery County Chest X-ray Datasets

The Montgomery dataset consists of 138 frontal CXRs (80 are normal cases, 58 are cases with manifestations of tuberculosis). The CXRs in this data set were acquired from the Tuberculosis Control Program of the Department of Healthand Human Services of Montgomery County, MD, USA. The Shenzhen dataset consists of 662 frontal chest X-rays (326 are normal cases, 336 are cases with manifestations of TB) that were collected as part of routine care at the Shenzhen No.3 Hospital in Shenzhen, China. We performed binary classification on this dataset, predicting whether a CXR contained manifestations of tuberculosis.

5. Pulmonary Edema Severity Grades Based on MIMIC-CXR

This dataset focuses on the pulmonary edema severity grades extracted from the MIMIC-CXR dataset through regular expression labeling from radiology reports, expert labeling from radiology reports and consensus labeling from chest radiographs. Any patient IDs that were repeated were only retained in consensus labeling. This dataset contains 12,022 images. The edema severity scores were defined as 0: none, 1: vascular congestion, 2: interstitial edema and 3: alveolar edema. We classified the subtlety of edema in each CXR in this dataset.

6. Indiana Chest X-ray Dataset

This dataset consists of CXR reports for posterior-anterior (PA) chest X-ray examinations from 2 large hospital systems within the Indiana Network for Patient Care database. The dataset contains 3850 chest X-ray reports and 7469 images. Most reports have corresponding frontal and lateral views. The entries under the ‘problems’ column of the dataset were separated into individual terms by splitting each entry on semicolons. Each ‘problem’ term was then set as a new column. We found a few terms that were identical except for differences in capitalization or spacing, so we added a new column named 'Thoracic Vertebrae Final' that represents both 'Thoracic Vertebrae’ and ‘Thoracic vertebrae' and a column named 'Subcutaneous Emphysema Final' that represents both ‘Subcutaneous  Emphysema' and 'Subcutaneous Emphysema'. All other problem terms were unique. Most terms applied to fewer than 1% of the chest X-ray reports, so we filtered the labels to preserve only those which were true for at least 1% of the reports. This filtration provided 20 labels. We perform 20 binary classification tasks on this dataset, predicting whether each of the 20 labels applies to a given CXR.

7. Japan Society for Radiology

The database includes 247 conventional chest radiographs obtained from 13 medical centers in Japan and one institution in the United States. Of the 154 chest X-rays containing nodules, 100 X-rays displayed malignant nodules and 54 displayed benign nodules. The sequence of lung nodule images is arranged according to the degree of subtlety from 5 (obvious) to 1 (extremely subtle). The degree of subtlety of the lung nodule scores were designated based on the consensus of three chest radiologists. The levels of subtlety in detecting a lung nodule were defined as follows: level 1, extremely subtle (where detection is extremely difficult because of low contrast, small size, or overlap with a normal structure); level 2, very subtle (where detection is very difficult); level 3, subtle (where detection is difficult); level 4, relatively obvious (where detection is relatively easy); and level 5, obvious (where detection is easy). We set subtlety scores to 0 to indicate absence of nodules. On this dataset, we predict the subtlety score for each chest X-ray image.

8. PadChest

This dataset includes more than 160,868 images obtained from 69,882 patients at Hospital San Juan Hospital (Spain) from 2009 to 2017. The images come with additional information on image acquisition and patient demography. The reports were labeled with 174 different radiographic findings, 19 differential diagnoses and 104 anatomic locations. Of these reports, 24.3% were manually annotated by trained physicians and the remaining set was labeled through supervised learning using a recurrent neural network with attention mechanisms. We restricted our analysis to only the physician-labeled CXRs and studied the following labels: Pneumonia, Atypical pneumonia, Tuberculosis, Tuberculosis sequelae, Lung metastasis, Lymphangitis carcinomatosa, Lepidic adenocarcinoma, Pulmonary fibrosis, Post radiotherapy changes, Asbestosis signs, Emphysema, COPD signs, Heart insufficiency, Respiratory distress, Pulmonary hypertension, Pulmonary artery hypertension, Pulmonary venous hypertension, Pulmonary edema and Bone metastasis. We perform differential diagnosis on this dataset, predicting which conditions are present in any given CXR.

9. REFLACX

The REFLACX dataset consists of CXRs from the MIMIC-CXR dataset, filtered to contain chest X-rays with only posterior-anterior views. Flipped or rotated CXRs with major parts of the lung missing from the field of view were excluded. We study the following 9 labels: Atelectasis, Consolidation, Enlarged cardiac silhouette, Groundglass opacity, High lung volume / emphysema, Lung nodule or mass, Pleural abnormality, Pneumothorax, and Pulmonary edema. We perform 9 binary classification tasks on this dataset, predicting whether each of these labels applies to any given CXR.

10. BRAX

The BRAX dataset contains 40,967 chest X-rays of 19,351 patients obtained from Hospital Israelita Albert Einstein in Brazil. This dataset includes 14 labels: Atelectasis, Cardiomegaly, Consolidation, Edema, Enlarged cardiomediastinum, Fracture, Lung lesion, Lung opacity, No findings, Pleural effusion, Pleural other, Pneumonia, Pneumothorax, and Support devices. Labels were extracted and translated to English from free-text Brazilian Portuguese radiology reports using natural language processing. Three Brazilian Portuguese native speaking radiologists verified the translations. The Chexpert Label Extraction Algorithm was then used to derive labels from either the findings section or the final section of the report (if neither impression nor findings sections were present). Each CXR was labeled as either “positive,” “negative” or “uncertain” for each of the conditions. We grouped “negative” and “uncertain” images together into one class and placed “positive” images into a second class. We performed 14 binary classification tasks on this dataset, predicting whether each CXR is positive for the 14 labels.

11. NIH ChestX-ray14

The NIH-released dataset ChestX-ray14 contains 112,120 frontal chest radiographs from 30,805 patients, mined from the NIH PACS system. Each chest X-ray has fourteen binary labels indicating the presence or absence of these pathologies: Atelectasis, Cardiomegaly, Effusion, Infiltration, Mass, Nodule, Pneumonia, Pneumothorax, Consolidation, Edema, Emphysema, Fibrosis, Pleural Thickening and Hernia. We performed 14 binary classification tasks on this dataset, predicting whether each of the 14 labels applies to a given CXR.

12. VinDr-CXR

The VinDr-CXR dataset, compiled between 2018 and 2020, consists of 20,925 chest X-rays from local PACS servers of Hospital 108 and the Hanoi Medical University Hospital, two of the largest hospitals in Vietnam. Images come from adult patients and are stored in DICOM format. The dataset contains labels for 28 findings and diagnoses. In the provided training set, each chest X-ray comes with annotations from three different radiologists and thus has three sets of 28 binary labels. We took the majority label (between the three labels for each of the 28 findings) for each chest X-ray, resulting in one set of 28 labels for each chest X-ray. Next, we filtered the findings to keep only those which occurred at least 10 times in the entire training set. We thus arrived at our final set of 18 labels, namely Aortic enlargement, Atelectasis, Calcification, Cardiomegaly, Consolidation, ILD, Infiltration, Lung Opacity, Nodule/Mass, Pleural effusion, Pleural thickening, Pulmonary fibrosis, Other lesion, Lung tumor, Pneumonia, Tuberculosis, Other diseases and No finding. Our test set contained the 3,000 annotated chest X-rays from the test set provided in the VinDr dataset. We did not have to take consensus for the test set, as this step had already been carried out by the VinDr-CXR dataset creators. We performed 18 binary classification tasks on this dataset, predicting whether each of the 18 labels applies to a given CXR

13. TB Portals

The version of the TB Portals dataset used here is a public dataset consisting of 1532 chest X-rays collected from medical centers and research studies of active TB disease in Eastern Europe and Central Asia (Azerbaijan, Belarus, Georgia, India, Kazakhstan, Moldova, Nigeria, Romania, Ukraine). It also contains patient-level metadata, including sputum microscopy for Acid Fast Bacilli (with grades ranging from negative to 4+ as detailed below) and timing of sputum testing and CXR. The dataset is collected by NIH/NIAID and is actively being expanded to other countries. We filtered the dataset to identify the chest X-rays taken closest to the first day of treatment. Next, we filtered the dataset to retain the earliest drug susceptibility test results that occurred within 60 days of the first day of treatment. Finally, we filtered out patient records with smear grades that did not match one of the labels ‘Negative’, '10 to 99 in 100 (1+)', '1 to 9 in 1 (2+)', '1 to 9 in 100 (1-9/100)', '10 to 99 in 1 (3+)' , and 'More than 99 in 1 (4+)'. We considered chest X-rays negative if they were labeled “Negative,” and positive otherwise. We performed binary classification on this dataset, predicting whether a chest X-ray had a positive or negative smear grade.

#### Supplementary note 2

##### Performance of self-supervised models across different clinical tasks

###### Multi-Label

We included 7 multi-label datasets in this study. RANZCR-CLiP addresses line and tube placements, while Indiana, PadChest, REFLACX, BRAX, NIH and VinDR focus on chest X-rays which may contain several pathologies at once. No single model achieved the highest AUC for all tasks on a single dataset.

On RANZCR-CLiP, all self-supervised models outperformed the ImageNet-pretrained model on all tasks, in both the finetuning and linear probing experiments. When finetuned, the best-performing model was GLoRIA, with an average AUC of 0.883 across all tasks. The ImageNet-pretrained model had an average AUC of 0.813. All self-supervised models had a high performance for Endotracheal Tube Assessment (Normal, Borderline and Abnormal), with the highest performance seen for normal ETT placement (AUC 0.984 with GLoRIA). For Nasogastric Tube (NGT) assessment, the highest performance was seen with normal placement. (AUC 0.963 with GLoRIA). There was an overall drop in performance for the assessment of central catheters (CVC), with the highest performance also achieved by GLoRIA detecting normal placement (AUC 0.803).
For linear probing, GLoRIA was also the highest performing model, with an average AUC of 0.859 across all tasks. For ETT assessment, the highest performance was again seen with normal placement (AUC 0.979 with ConVIRT). For NGT placement, we again saw the best performance with normal placement (AUC 0.941 with GLoRIA). For CVC, the highest performance was seen instead for abnormal placement (AUC 0.696 with GLoRIA); we again see the same drop in performance as with finetuning. Refer to Table 6 for more details.

On the Indiana dataset, the highest AUC was achieved by CXR-pretrained models on 20 of the 21 labels assessed. After full finetuning, REFERS was on average the best-performing model across tasks (AUC 0.799), while the ImageNet-pretrained model had an AUC of only 0.727. However, ConVIRT was the top-performing model on each of the three tasks with highest performance (pleural effusion AUC 0.946, edema AUC 0.938, and cardiomegaly AUC 0.926). With linear probing, the best-performing model on average across tasks was ConVIRT (AUC 0.822), outperforming all finetuned models. On the three tasks with highest performance, ConVIRT was again the best-performing model (edema AUC 0.979, pleural effusion AUC 0.950, and pulmonary congestion AUC 0.932). Refer to Table 9 for more details.

On PadChest, REFERS had the highest performance after full finetuning (AUC 0.646), while the ImageNet-pretrained model had an AUC of 0.635. When models were finetuned, the tasks with the highest performance were pulmonary edema (AUC 0.777 with MoCo-CXR), heart insufficiency (AUC 0.771 with S2MTS2) and pneumonia (AUC 0.692 with CXR-RePaiR-CLIP). With linear probing, no self-supervised models outperformed the finetuned ImageNet-pretrained model (AUC 0.635), but CXR-RePaiR-CLIP nearly matched its performance (AUC 0.631). We again saw the highest performance on the same tasks: pulmonary edema with the pretrained ImageNet model (AUC 0.752 after finetuning), heart insufficiency with GLoRIA (AUC 0.710) and pneumonia detection with CXR-RePaiR-CLIP (AUC 0.686). Refer to Table 7 for more details.

On REFLACX, a self-supervised model outperformed the ImageNet-pretrained on every task, both when we finetuned models and when we performed linear probing. On average across tasks, the best-performing model after finetuning was ConVIRT (AUC 0.847), while the ImageNet-pretrained model had an AUC of only 0.744. When breaking down results by task, finetuned models achieved the highest performance on pleural abnormality (AUC of 0.925 with both ConVIRT and GLoRIA), cardiomegaly with ConVIRT (AUC 0.922) and emphysema with S2MTS2 (AUC 0.918). When performing linear probing, the best performing model was GLoRIA (AUC 0.853), and the tasks with the highest performance were again emphysema (AUC 0.950 with REFERS), pleural abnormality (AUC 0.925 with GloRIA) and cardiomegaly (AUC 0.918 with ConVIRT). Refer to Table 11 for more details.

On BRAX, self-supervised models outperformed the supervised model on 13 of 14 labels after finetuning. Perhaps more importantly, they outperformed the supervised model on all 14 labels with linear probing. On average across tasks, the best-performing finetuned model was ConVIRT (AUC 0.820), while the ImageNet-pretrained model achieved only an AUC of 0.779. On specific tasks, we saw the highest performance on pleural effusion detection (AUC 0.929), pneumothorax (AUC 0.926), and cardiomegaly (AUC 0.918), all achieved by ConVIRT. With linear probing, ConVIRT was also the highest performing model on average across tasks (AUC 0.821), and the same three tasks elicited the highest performance, again with ConVIRT: pneumothorax detection, (AUC 0.942), pleural effusion (AUC 0.930) and cardiomegaly (AUC 0.916). Refer to Table 12 for more details.

On the NIH dataset, the highest performance on each task was achieved by a self-supervised model, both for finetuning and linear probing. Among finetuned models, ConVIRT was the best performing model on average across tasks (AUC 0.813), while the ImageNet-pretrained model had an AUC of 0.706. The highest per-task performance was achieved when detecting cardiomegaly (AUC 0.925), pleural thickening (AUC 0.910) and pleural effusion (AUC 0.899); again ConVIRT was the best model on each of these tasks. After linear probing, the best performing model was REFERS (AUC 0.797). The highest per-task performance was also achieved on pleural thickening (REFERS with AUC of 0.908), cardiomegaly (GLoRIA with AUC of 0.899) and pleural effusion (REFERS with AUC of 0.890). Refer to Table 13 for more details.

On VinDR, the highest performance on each task was consistently achieved by a self-supervised model, both for finetuning and linear probing. After finetuning, the highest average performance across tasks was seen with REFERS (AUC 0.879), while the ImageNet-pretrained model had an AUC of 0.814. The best per-task results were also seen with REFERS on pleural effusion detection (AUC 0.952), cardiomegaly (AUC 0.935) and no finding (AUC 0.916). When trained with linear probing, REFERS achieved an even higher performance (AUC 0.878) than when finetuned. The best per-task results were also seen with REFERS on pleural effusion detection (AUC 0.955), cardiomegaly (AUC 0.948) and pneumonia (AUC 0.933). Refer to Table 14 for more details.

Multi-Class

We evaluated two multi-class datasets, MIMIC-CXR for edema severity classification and JSRT for subtlety of lung nodules. Performance for all models was lower overall in comparison to other tasks and datasets.

On the JSRT dataset, MoCo-CXR was the best-performing finetuned model on average across classes (AUC 0.558) and GLoRIA was the best-performing model after linear probing (AUC 0.587). With finetuning, the highest AUC for a subtlety score of 0 was achieved by GLoRIA (AUC 0.757). The ImageNet-pretrained model had the highest AUC for a subtlety score of 5 (AUC 0.716). For subtleties of 1 through 4, the best performance was below 0.650 for all models including ImageNet pretrained models.

When performing linear probing, the highest single-class AUC was for a subtlety score of 5 with GLoRIA (AUC 0.755). The second highest single-class AUC was seen here for a subtlety score of 2 with MedAug (AUC 0.735). For a subtlety score of 0, CXR-RePaiR-CLIP had the best performance with an AUC of 0.698. Refer to Table 10 for more details.

On the MIMIC-CXR dataset, ConVIRT was the best-performing model (finetuning average AUC 0.802, linear probing AUC 0.791); the highest AUC was seen for sevrerity of 0 (AUC of 0.864 after finetuning and 0.849 after linear probing) followed by severity of 3 (AUC of 0.847 after finetuning and 0.839 ater linear probing.) For severities of 1 and 2, performance for the best-performing model (ConVIRT) were between 0.702 and 0.792 AUC with both finetuning and linear probing. Refer to Table 8 for more details.

Binary Classification

We evaluated 4 datasets with binary classification tasks: SIIM-ACR for pneumothorax detection, Pediatric Pneumonia for pneumonia detection, Shenzhen and Montgomery for Tuberculosis detection and the TB Portals dataset for predicting smear positivity based on chest-X-rays. Because Shenzhen and Montgomery are particularly small datasets, we combined them for training and validation. When detecting the presence of a single pathology, all models demonstrated excellent performance.

On the SIIM-ACR dataset, GLoRIA had the best performance after both finetuning and linear probing (AUC 0.934 and 0.919 respectively.) For comparison, the ImageNet-pretrained model had an AUC of 0.917. On the Pediatric Pneumonia dataset, GLoRIA achieved the best performance with finetuning (AUC 0.983), while ConVIRT performed best after linear probing (AUC 0.993). The ImageNet-pretrained model had an AUC of 0.930. On the Shenzhen and Montgomery dataset, REFERS had the best performance with finetuning (AUC 0.973), while ConVIRT was again the best performer after linear probing (AUC 0.955). The ImageNet supervised model had an AUC of 0.923. The TB Portals dataset is also binary (positive or negative); however, the task was to predict smear positivity of sputum based on the chest-X-ray. On this dataset, performance was the lowest of any binary task. The highest performance was seen with REFERS with both finetuning and linear probing (AUC 0.714 and 0.715 respectively). The pretrained ImageNet model had an AUC of 0.707. For more details of these results see Table 4 (finetuning results) and Table 5 (linear probing results).

#### Supplementary note 3

| FINETUNING | ImageNet-Pretrained Baseline | MedAug | CXR-RePaiR-CLIP | REFERS | ConVIRT | GLoRIA | S2MTS2 | MoCo-CXR |
|---|---|---|---|---|---|---|---|---|
| RANZCR CLiP | 0.813 [0.791, 0.833] | 0.824 [0.798, 0.845] | 0.819 [0.797, 0.840] | 0.857 [0.839, 0.875] | 0.876 [0.861, 0.893] | **0.883 [0.865, 0.900]** | 0.840 [0.819, 0.860] | 0.832 [0.804, 0.859] |
| SIIM-ACR Pneumothorax | 0.917 [0.905, 0.929] | 0.903 [0.890, 0.915] | 0.827 [0.806, 0.842] | 0.921 [0.910, 0.933] | 0.925 [0.918, 0.937] | **0.934 [0.924, 0.946]** | 0.918 [0.908, 0.926] | 0.912 [0.900, 0.924] |
| Pediatric Pneumonia | 0.930 [0.898, 0.953] | 0.967 [0.951, 0.983] | 0.953 [0.932, 0.965] | 0.978 [0.965, 0.988] | 0.978 [0.964, 0.989] | **0.983 [0.971, 0.992]** | 0.973 [0.961, 0.985] | 0.951 [0.927, 0.971] |
| Shenzhen + Montgomery | 0.923 [0.876, 0.967] | 0.920 [0.867, 0.966] | 0.882 [0.830, 0.937] | **0.973 [0.949, 0.992]** | 0.964 [0.937, 0.990] | 0.969 [0.942, 0.990] | 0.917 [0.885, 0.958] | 0.929 [0.891, 0.964] |
| Pulmonary Edema Severity | 0.713 [0.685, 0.740] | 0.753 [0.728, 0.776] | 0.665 [0.633, 0.694] | 0.789 [0.770, 0.806] | **0.802 [0.779, 0.821]** | 0.755 [0.733, 0.774] | 0.730 [0.704, 0.754] | 0.736 [0.712, 0.761] |
| TB Portals | 0.707 [0.665, 0.766] | 0.681 [0.615, 0.7527] | 0.685 [0.637, 0.746] | **0.714 [0.658, 0.775]** | 0.679 [0.630, 0.733] | 0.683 [0.631, 0.746] | 0.698 [0.655, 0.761] | 0.695 [0.6425, 0.758] |
| Indiana University | 0.727 [0.650, 0.798] | 0.733 [0.664, 0.795] | 0.732 [0.658, 0.795] | **0.799 [0.736, 0.866]** | 0.793 [0.729, 0.851] | 0.739 [0.668, 0.803] | 0.767 [0.691, 0.829] | 0.718 [0.643, 0.782] |
| JSRT | 0.543 [0.347, 0.709] | 0.542 [0.368, 0.715] | 0.511 [0.362, 0.655] | 0.542 [0.366, 0.704] | 0.555 [0.424, 0.686] | 0.554 [0.417, 0.696] | 0.489 [0.350, 0.635] | **0.558 [0.388, 0.714]** |
| PadChest | 0.635 [0.573, 0.694] | 0.582 [0.520, 0.645] | 0.645 [0.581, 0.707] | 0.646 [0.571, 0.702] | 0.621 [0.559, 0.681] | 0.619 [0.549, 0.677] | **0.659 [0.590, 0.718]** | 0.635 [0.573, 0.698] |
| REFLACX | 0.744 [0.674, 0.804] | 0.747 [0.681, 0.810] | 0.811 [0.747, 0.867] | 0.825 [0.777, 0.884] | **0.847 [0.793, 0.903]** | 0.843 [0.788, 0.893] | 0.792 [0.727, 0.855] | 0.744 [0.678, 0.805] |
| BRAX | 0.779 [0.734, 0.821] | 0.787 [0.745, 0.830] | 0.792 [0.743, 0.838] | 0.808 [0.766, 0.850] | **0.820 [0.780, 0.861]** | 0.796 [0.752, 0.837] | 0.801 [0.757, 0.844] | 0.779 [0.736, 0.820] |
| NIH ChestXRay14 | 0.706 [0.683, 0.731] | 0.716 [0.692, 0.743] | 0.764 [0.741, 0.790] | 0.798 [0.775, 0.822] | **0.813 [0.793, 0.834]** | 0.795 [0.765, 0.819] | 0.771 [0.745, 0.791] | 0.707 [0.682, 0.733] |
| VinDr-CXR | 0.814 [0.780, 0.847] | 0.794 [0.763, 0.826] | 0.840 [0.811, 0.868] | **0.879 [0.850, 0.904]** | 0.859 [0.832, 0.886] | 0.836 [0.807, 0.863] | 0.799 [0.763, 0.833] | 0.801 [0.768, 0.832] |
| Average Peformance Across Datasets | 0.765 | 0.765 | 0.764 | 0.810 | 0.810 | 0.799 | 0.781 | 0.769 |

**Table 4. Finetuning results -** Average performance (AUC) across all labels per dataset of an ImageNet pretrained model  and seven self-supervised CXR pretrained models. The best performance per dataset is indicated in bold.

| LINEAR PROBING | ImageNet-Pretrained Baseline | MedAug | CXR-RePaiR-CLIP | REFERS | ConVIRT | GLoRIA | S2MTS2 | MoCo-CXR |
|---|---|---|---|---|---|---|---|---|
| RANZCR CLiP | 0.813 [0.791, 0.833] | 0.789 [0.766, 0.814] | 0.816 [0.796, 0.837] | 0.837 [0.818, 0.855] | 0.849 [0.833, 0.865] | **0.859 [0.840, 0.875]** | 0.788 [0.766, 0.811] | 0.801 [0.779, 0.826] |
| SIIM-ACR Pneumothorax | 0.917 [0.905, 0.929] | 0.810 [0.793, 0.828] | 0.876 [0.865, 0.892] | 0.900 [0.886, 0.910] | 0.909 [0.899, 0.919] | **0.919 [0.908, 0.928]** | 0.832 [0.816, 0.846] | 0.811 [0.797, 0.829] |
| Pediatric Pneumonia | 0.930 [0.898, 0.953] | 0.942 [0.927, 0.959] | 0.974 [0.9617, 0.984] | 0.985 [0.976, 0.994] | **0.993 [0.988, 0.999]** | 0.989 [0.979, 0.996] | 0.851 [0.815, 0.883] | 0.696 [0.661, 0.735] |
| Shenzhen + Montgomery | 0.923 [0.876, 0.967] | 0.826 [0.766, 0.887] | 0.894 [0.839, 0.940] | 0.933 [0.873, 0.958] | **0.955 [0.916, 0.985]** | 0.944 [0.907, 0.976] | 0.869 [0.826, 0.917] | 0.892 [0.838, 0.943] |
| Pulmonary Edema Severity | 0.713 [0.685, 0.740] | 0.698 [0.671, 0.723] | 0.774 [0.753, 0.797] | 0.773 [0.747, 0.795] | **0.791 [0.772, 0.812]** | 0.753 [0.728, 0.777] | 0.687 [0.660, 0.714] | 0.721 [0.692, 0.746] |
| TB Portals | 0.707 [0.665, 0.766] | 0.652 [0.601, 0.722] | 0.702 [0.643, 0.760] | **0.715 [0.652, 0.772]** | 0.685 [0.627, 0.739] | 0.712 [0.660, 0.783] | 0.691 [0.635, 0.745] | 0.693 [0.638, 0.758] |
| Indiana University | 0.727 [0.650, 0.798] | 0.759 [0.689, 0.818] | 0.783 [0.716, 0.839] | 0.809 [0.752, 0.862] | **0.822 [0.759, 0.875]** | 0.751 [0.676, 0.824] | 0.740 [0.664, 0.809] | 0.728 [0.656, 0.798] |
| JSRT | 0.543 [0.347, 0.709] | 0.575 [0.425, 0.731] | 0.522 [0.368, 0.682] | 0.551 [0.395, 0.705] | 0.520 [0.381, 0.669] | **0.587 [0.419, 0.747]** | 0.444 [0.289, 0.595] | 0.527 [0.357, 0.685] |
| PadChest | **0.635 [0.573, 0.694]** | 0.583 [0.517, 0.653] | 0.631 [0.558, 0.692] | 0.596 [0.521, 0.659] | 0.602 [0.533, 0.667] | 0.621 [0.546, 0.686] | 0.591 [0.527, 0.653] | 0.606 [0.543, 0.661] |
| REFLACX | 0.744 [0.674, 0.804] | 0.773 [0.709, 0.829] | 0.831 [0.771, 0.889] | 0.845 [0.792, 0.894] | 0.846 [0.785, 0.900] | **0.853 [0.800, 0.903]** | 0.774 [0.711, 0.837] | 0.750 [0.688, 0.813] |
| BRAX | 0.779 [0.734, 0.821] | 0.776 [0.734, 0.820] | 0.799 [0.758, 0.838] | 0.803 [0.765, 0.845] | **0.821 [0.783, 0.859]** | 0.793 [0.746, 0.834] | 0.767 [0.720, 0.815] | 0.765 [0.717, 0.815] |
| NIH ChestXRay14 | 0.706 [0.683, 0.731] | 0.614 [0.586, 0.641] | 0.788 [0.765, 0.810] | **0.797 [0.772, 0.822]** | 0.743 [0.717, 0.768] | 0.723 [0.697, 0.752] | 0.587 [0.563, 0.618] | 0.640 [0.613, 0.670] |
| VinDr-CXR | 0.814 [0.780, 0.847] | 0.615 [0.572, 0.662] | 0.862 [0.833, 0.890] | **0.878 [0.851, 0.904]** | 0.758 [0.721, 0.799] | 0.675 [0.635, 0.717] | 0.636 [0.584, 0.688] | 0.602 [0.550, 0.654] |
| Average Peformance Across Datasets | 0.765 | 0.724 | 0.789 | 0.802 | 0.792 | 0.783 | 0.712 | 0.710 |

**Table 5. Linear Probing results -** Average performance (AUC) across all labels per dataset of an ImageNet pretrained model (with Finetuning) and seven self-supervised CXR pretrained models. (Linear Probing). The best performance per dataset is indicated in bold.

| FINETUNING | ImageNet-Pretrained Baseline | MedAug | CXR-RePaiR-CLIP | REFERS | ConVIRT | GLoRIA | S2MTS2 | MoCo-CXR |
|---|---|---|---|---|---|---|---|---|
| ETT - Abnormal | 0.867 [0.814, 0.915] | 0.833 [0.744, 0.897] | 0.874 [0.816, 0.922] | 0.919 [0.869, 0.960] | **0.941 [0.915, 0.970]** | 0.899 [0.832, 0.951] | 0.895 [0.846, 0.934] | 0.819 [0.722, 0.930] |
| ETT - Borderline | 0.891 [0.881, 0.904] | 0.897 [0.884, 0.910] | 0.904 [0.893, 0.916] | 0.904 [0.887, 0.919] | 0.930 [0.922, 0.940] | **0.933 [0.924, 0.942]** | 0.907 [0.899, 0.916] | 0.873 [0.854, 0.893] |
| ETT - Normal | 0.964 [0.959, 0.968] | 0.978 [0.975, 0.981] | 0.970 [0.966, 0.974] | 0.980 [0.977, 0.983] | 0.983 [0.980, 0.986] | **0.984 [0.982, 0.987]** | 0.978 [0.974, 0.982] | 0.978 [0.975, 0.982] |
| NGT - Abnormal | 0.807 [0.769, 0.850] | 0.793 [0.745, 0.837] | 0.780 [0.728, 0.826] | 0.870 [0.835, 0.908] | 0.829 [0.786, 0.882] | **0.843 [0.802, 0.879]** | 0.832 [0.787, 0.878] | 0.815 [0.756, 0.873] |
| NGT - Borderline | 0.801 [0.760, 0.831] | 0.773 [0.730, 0.803] | 0.803 [0.762, 0.837] | **0.860 [0.833, 0.889]** | 0.847 [0.823, 0.874] | 0.851 [0.827, 0.881] | 0.829 [0.781, 0.866] | 0.790 [0.759, 0.818] |
| NGT - Incompletely Imaged | 0.931 [0.922, 0.941] | 0.945 [0.934, 0.955] | 0.936 [0.925, 0.945] | 0.944 [0.936, 0.953] | 0.947 [0.939, 0.955] | **0.954 [0.948, 0.960]** | 0.942 [0.932, 0.950] | 0.923 [0.912, 0.936] |
| NGT - Normal | 0.928 [0.918, 0.935] | 0.940 [0.933, 0.947] | 0.925 [0.918, 0.933] | 0.942 [0.937, 0.948] | 0.957 [0.952, 0.961] | **0.963 [0.960, 0.967]** | 0.935 [0.926, 0.943] | 0.936 [0.930, 0.942] |
| CVC - Abnormal | 0.632 [0.613, 0.654] | 0.663 [0.646, 0.680] | 0.654 [0.630, 0.676] | 0.695 [0.674, 0.717] | 0.734 [0.709, 0.757] | **0.756 [0.735, 0.778]** | 0.663 [0.644, 0.687] | 0.679 [0.653, 0.700] |
| CVC - Borderline | 0.604 [0.588, 0.617] | 0.648 [0.632, 0.663] | 0.604 [0.588, 0.619] | 0.655 [0.640, 0.670] | 0.718 [0.704, 0.733] | **0.732 [0.717, 0.747]** | 0.633 [0.617, 0.655] | 0.672 [0.655, 0.686] |
| CVC - Normal | 0.604 [0.592, 0.619] | 0.660 [0.646, 0.676] | 0.603 [0.591, 0.621] | 0.668 [0.651, 0.681] | 0.754 [0.740, 0.770] | **0.803 [0.793, 0.814]** | 0.634 [0.619, 0.651] | 0.708 [0.694, 0.726] |
| Swan Ganz Catheter Present | 0.910 [0.888, 0.928] | 0.930 [0.913, 0.945] | 0.960 [0.949, 0.970] | 0.994 [0.992, 0.997] | 0.999 [0.998, 1] | **0.999 [0.999, 0.999]** | 0.994 [0.988, 0.997] | 0.954 [0.934, 0.965] |
| Overall AUC | 0.813 [0.791, 0.833] | 0.824 [0.798, 0.845] | 0.819 [0.797, 0.840] | 0.857 [0.839, 0.875] | 0.876 [0.861, 0.893] | **0.883 [0.865, 0.900]** | 0.840 [0.819, 0.860] | 0.832 [0.804, 0.859] |

| LINEAR PROBING | ImageNet-Pretrained Baseline | MedAug | CXR-RePaiR-CLIP | REFERS | ConVIRT | GLoRIA | S2MTS2 | MoCo-CXR |
|---|---|---|---|---|---|---|---|---|
| ETT - Abnormal | 0.867 [0.814, 0.915] | 0.802 [0.734, 0.869] | 0.892 [0.857, 0.930] | **0.929 [0.896, 0.959]** | 0.913 [0.887, 0.942] | 0.898 [0.840, 0.934] | 0.851 [0.798, 0.912] | 0.849 [0.812, 0.920] |
| ETT - Borderline | 0.891 [0.881, 0.904] | 0.860 [0.843, 0.874] | 0.900 [0.889, 0.911] | 0.908 [0.897, 0.919] | **0.922 [0.911, 0.930]** | 0.920 [0.908, 0.928] | 0.853 [0.833, 0.868] | 0.878 [0.862, 0.892] |
| ETT - Normal | 0.964 [0.959, 0.968] | 0.926 [0.920, 0.933] | 0.951 [0.946, 0.956] | 0.973 [0.970, 0.977] | **0.979 [0.976, 0.982]** | 0.978 [0.975, 0.982] | 0.897 [0.889, 0.907] | 0.926 [0.920, 0.934] |
| NGT - Abnormal | 0.807 [0.769, 0.850] | 0.785 [0.732, 0.843] | 0.801 [0.756, 0.855] | 0.813 [0.767, 0.869] | 0.824 [0.779, 0.868] | **0.864 [0.818, 0.899]** | 0.793 [0.745, 0.852] | 0.792 [0.734, 0.852] |
| NGT - Borderline | 0.801 [0.760, 0.831] | 0.798 [0.762, 0.827] | 0.792 [0.748, 0.829] | 0.835 [0.801, 0.862] | 0.856 [0.835, 0.884] | **0.860 [0.836, 0.883]** | 0.758 [0.726, 0.787] | 0.775 [0.734, 0.811] |
| NGT - Incompletely Imaged | 0.931 [0.922, 0.941] | 0.885 [0.875, 0.899] | 0.925 [0.914, 0.934] | 0.926 [0.915, 0.935] | 0.939 [0.932, 0.947] | **0.941 [0.931, 0.948]** | 0.878 [0.865, 0.890] | 0.906 [0.895, 0.917] |
| NGT - Normal | 0.928 [0.918, 0.935] | 0.886 [0.875, 0.896] | 0.914 [0.906, 0.924] | 0.920 [0.913, 0.929] | 0.937 [0.930, 0.944] | **0.942 [0.935, 0.950]** | 0.862 [0.854, 0.872] | 0.894 [0.886, 0.903] |
| CVC - Abnormal | 0.632 [0.613, 0.654] | 0.634 [0.620, 0.660] | 0.641 [0.619, 0.664] | 0.653 [0.630, 0.675] | 0.670 [0.646, 0.691] | **0.696 [0.674, 0.716]** | 0.637 [0.620, 0.656] | 0.646 [0.621, 0.670] |
| CVC - Borderline | 0.604 [0.588, 0.617] | 0.606 [0.593, 0.624] | 0.612 [0.599, 0.626] | 0.640 [0.625, 0.653] | 0.655 [0.640, 0.670] | **0.671 [0.658, 0.688]** | 0.617 [0.601, 0.632] | 0.607 [0.595, 0.622] |
| CVC - Normal | 0.604 [0.592, 0.619] | 0.583 [0.570, 0.599] | 0.607 [0.591, 0.625] | 0.629 [0.616, 0.642] | 0.653 [0.638, 0.666] | **0.688 [0.673, 0.704]** | 0.593 [0.578, 0.608] | 0.591 [0.578, 0.605] |
| Swan Ganz Catheter Present | 0.910 [0.888, 0.928] | 0.912 [0.900, 0.926] | 0.943 [0.931, 0.955] | 0.977 [0.969, 0.986] | 0.992 [0.989, 0.995] | **0.996 [0.993, 0.998]** | 0.927 [0.913, 0.941] | 0.944 [0.930, 0.958] |
| Overall AUC | 0.813 [0.791, 0.833] | 0.789 [0.766, 0.814] | 0.816 [0.796, 0.837] | 0.837 [0.818, 0.855] | 0.849 [0.833, 0.865] | **0.859 [0.840, 0.875]** | 0.788 [0.766, 0.811] | 0.801 [0.779, 0.826] |

**Table 6. RANZCR CLiP Results -** AUC across all set of labels for Finetuning and Linear Probing.

| FINETUNING | ImageNet-Pretrained Baseline | MedAug | CXR-RePaiR-CLIP | REFERS | ConVIRT | GLoRIA | S2MTS2 | MoCo-CXR |
|---|---|---|---|---|---|---|---|---|
| **Lung Metastasis** | 0.481 [0.378, 0.590] | 0.593 [0.465, 0.695] | 0.565 [0.450, 0.681] | **0.6338 [0.530, 0.728]** | 0.557 [0.451, 0.648] | 0.507 [0.397, 0.610] | 0.581 [0.476, 0.678] | 0.583 [0.470, 0.687] |
| **Tuberculosis Sequelae** | **0.610 [0.507, 0.722]** | 0.562 [0.501, 0.625] | 0.539 [0.448, 0.622] | 0.535 [0.451, 0.603] | 0.589 [0.501, 0.707] | 0.571 [0.451, 0.647] | 0.539 [0.418, 0.628] | 0.571 [0.455, 0.660] |
| **Emphysema** | 0.644 [0.585, 0.686] | 0.608 [0.543, 0.670] | 0.589 [0.534, 0.651] | 0.644 [0.549, 0.695] | 0.576 [0.517, 0.624] | 0.556 [0.482, 0.622] | **0.651 [0.589, 0.714]** | 0.623 [0.564, 0.687] |
| **Heart Insufficiency** | 0.702 [0.656, 0.747] | 0.552 [0.504, 0.609] | 0.745 [0.700, 0.791] | 0.755 [0.703, 0.802] | 0.668 [0.633, 0.700] | 0.702 [0.656, 0.740] | **0.771 [0.723, 0.812]** | 0.673 [0.631, 0.732] |
| **COPD Signs** | 0.587 [0.563, 0.601] | 0.558 [0.538, 0.582] | **0.614 [0.591, 0.635]** | 0.606 [0.584, 0.623] | 0.587 [0.565, 0.603] | 0.599 [0.577, 0.618] | 0.614 [0.591, 0.634] | 0.594 [0.570, 0.614] |
| **Normal** | 0.670 [0.657, 0.680] | 0.574 [0.558, 0.588] | 0.667 [0.653, 0.676] | 0.661 [0.647, 0.673] | 0.654 [0.641, 0.663] | 0.676 [0.661, 0.686] | 0.683 [0.668, 0.693] | **0.685 [0.673, 0.697]** |
| **Pulmonary Edema** | 0.752 [0.683, 0.822] | 0.635 [0.549, 0.731] | 0.777 [0.683, 0.858] | 0.705 [0.592, 0.785] | 0.715 [0.600, 0.823] | 0.744 [0.677, 0.824] | 0.738 [0.626, 0.822] | **0.777 [0.702, 0.853]** |
| **Pneumonia** | 0.677 [0.651, 0.706] | 0.630 [0.607, 0.658] | **0.692 [0.665, 0.723]** | 0.670 [0.635, 0.697] | 0.641 [0.615, 0.667] | 0.658 [0.633, 0.683] | 0.667 [0.632, 0.697] | 0.664 [0.636, 0.694] |
| **Pulmonary Fibrosis** | 0.595 [0.478, 0.696] | 0.522 [0.411, 0.646] | 0.615 [0.507, 0.728] | 0.604 [0.444, 0.709] | 0.603 [0.504, 0.690] | 0.558 [0.411, 0.666] | **0.687 [0.591, 0.786]** | 0.549 [0.452, 0.655] |
| **Overall AUC** | 0.635 [0.573, 0.694] | 0.582 [0.520, 0.645] | 0.645 [0.581, 0.707] | **0.646 [0.571, 0.702]** | 0.621 [0.559, 0.681] | 0.619 [0.549, 0.677] | 0.659 [0.590, 0.718] | 0.635 [0.573, 0.698] |

| LINEAR PROBING | ImageNet-Pretrained Baseline | MedAug | CXR-RePaiR-CLIP | REFERS | ConVIRT | GLoRIA | S2MTS2 | MoCo-CXR |
|---|---|---|---|---|---|---|---|---|
| **Lung Metastasis** | 0.481 [0.378, 0.590] | 0.555 [0.437, 0.681] | **0.596 [0.471, 0.683]** | 0.549 [0.440, 0.668] | 0.531 [0.432, 0.633] | 0.568 [0.453, 0.681] | 0.554 [0.432, 0.681] | 0.590 [0.488, 0.674] |
| **Tuberculosis Sequelae** | **0.610 [0.507, 0.722]** | 0.547 [0.469, 0.627] | 0.569 [0.472, 0.646] | 0.541 [0.453, 0.622] | 0.562 [0.469, 0.679] | 0.580 [0.490, 0.656] | 0.554 [0.464, 0.641] | 0.542 [0.466, 0.610] |
| **Emphysema** | **0.644 [0.585, 0.686]** | 0.565 [0.503, 0.636] | 0.618 [0.547, 0.680] | 0.621 [0.557, 0.683] | 0.605 [0.551, 0.661] | 0.613 [0.527, 0.672] | 0.597 [0.527, 0.649] | 0.584 [0.514, 0.642] |
| **Heart Insufficiency** | 0.702 [0.656, 0.747] | 0.617 [0.568, 0.669] | 0.695 [0.653, 0.747] | 0.658 [0.612, 0.697] | 0.643 [0.591, 0.702] | **0.710 [0.646, 0.754]** | 0.680 [0.647, 0.731] | 0.700 [0.658, 0.742] |
| **COPD Signs** | 0.587 [0.563, 0.601] | 0.565 [0.546, 0.582] | 0.602 [0.583, 0.621] | 0.578 [0.555, 0.597] | 0.578 [0.558, 0.600] | **0.604 [0.585, 0.622]** | 0.565 [0.546, 0.582] | 0.585 [0.568, 0.603] |
| **Normal** | **0.670 [0.657, 0.680]** | 0.601 [0.587, 0.612] | 0.638 [0.626, 0.652] | 0.608 [0.595, 0.625] | 0.630 [0.617, 0.642] | 0.669 [0.656, 0.680] | 0.619 [0.605, 0.633] | 0.633 [0.621, 0.645] |
| **Pulmonary Edema** | **0.752 [0.683, 0.822]** | 0.585 [0.470, 0.716] | 0.638 [0.500, 0.733] | 0.593 [0.457, 0.690] | 0.611 [0.477, 0.692] | 0.620 [0.502, 0.739] | 0.551 [0.450, 0.645] | 0.589 [0.463, 0.675] |
| **Pneumonia** | 0.677 [0.651, 0.706] | 0.640 [0.615, 0.670] | **0.686 [0.660, 0.717]** | 0.663 [0.641, 0.692] | 0.665 [0.643, 0.694] | 0.685 [0.660, 0.712] | 0.663 [0.636, 0.688] | 0.650 [0.619, 0.678] |
| **Pulmonary Fibrosis** | 0.595 [0.478, 0.696] | 0.576 [0.458, 0.684] | **0.633 [0.510, 0.753]** | 0.552 [0.379, 0.661] | 0.592 [0.460, 0.700] | 0.543 [0.398, 0.658] | 0.534 [0.435, 0.628] | 0.583 [0.490, 0.681] |
| **Overall AUC** | **0.635 [0.573, 0.694]** | 0.583 [0.517, 0.653] | 0.631 [0.558, 0.692] | 0.596 [0.521, 0.659] | 0.602 [0.533, 0.667] | 0.621 [0.546, 0.686] | 0.591 [0.527, 0.653] | 0.606 [0.543, 0.661] |

**Table 7. Padchest Results -** AUC across all diagnoses for Finetuning and Linear Probing.

| FINETUNING | ImageNet-Pretrained Baseline | MedAug | CXR-RePaiR-CLIP | REFERS | ConVIRT | GLoRIA | S2MTS2 | MoCo-CXR |
|---|---|---|---|---|---|---|---|---|
| Severity of 0 | 0.797 [0.783, 0.816] | 0.822 [0.809, 0.836] | 0.710 [0.697, 0.730] | 0.839 [0.826, 0.852] | **0.864 [0.852, 0.876]** | 0.821 [0.808, 0.837] | 0.805 [0.793, 0.818] | 0.807 [0.791, 0.824] |
| Severity of 1 | 0.635 [0.610, 0.668] | 0.6448 [0.622, 0.671] | 0.632 [0.602, 0.660] | 0.693 [0.673, 0.710] | **0.705 [0.677, 0.727]** | 0.640 [0.617, 0.667] | 0.661 [0.639, 0.687] | 0.637 [0.617, 0.671] |
| Severity of 2 | 0.678 [0.648, 0.700] | 0.752 [0.727, 0.777] | 0.613 [0.585, 0.634] | 0.778 [0.758, 0.794] | **0.793 [0.769, 0.810]** | 0.748 [0.726, 0.765] | 0.689 [0.659, 0.713] | 0.724 [0.703, 0.747] |
| Severity of 3 | 0.743 [0.698, 0.775] | 0.794 [0.755, 0.820] | 0.707 [0.650, 0.753] | 0.848 [0.823, 0.870] | **0.848 [0.819, 0.872]** | 0.810 [0.783, 0.828] | 0.766 [0.724, 0.797] | 0.777 [0.738, 0.803] |
| Averaged AUC | 0.713 [0.685, 0.740] | 0.753 [0.728, 0.776] | 0.665 [0.633, 0.694] | 0.789 [0.770, 0.806] | **0.802 [0.779, 0.821]** | 0.755 [0.733, 0.774] | 0.730 [0.704, 0.754] | 0.736 [0.712, 0.761] |

| LINEAR PROBING | ImageNet-Pretrained Baseline | MedAug | CXR-RePaiR-CLIP | REFERS | ConVIRT | GLoRIA | S2MTS2 | MoCo-CXR |
|---|---|---|---|---|---|---|---|---|
| Severity of 0 | 0.797 [0.783, 0.816] | 0.749 [0.726, 0.765] | 0.826 [0.812, 0.841] | 0.830 [0.809, 0.843] | **0.849 [0.838, 0.864]** | 0.804 [0.7874, 0.820] | 0.750 [0.730, 0.766] | 0.781 [0.763, 0.798] |
| Severity of 1 | 0.635 [0.610, 0.668] | 0.652 [0.628, 0.680] | 0.699 [0.676, 0.723] | 0.670 [0.645, 0.694] | **0.706 [0.684, 0.730]** | 0.663 [0.642, 0.688] | 0.626 [0.598, 0.652] | 0.642 [0.620, 0.660] |
| Severity of 2 | 0.678 [0.648, 0.700] | 0.655 [0.632, 0.679] | 0.751 [0.731, 0.774] | 0.764 [0.738, 0.783] | **0.772 [0.755, 0.793]** | 0.744 [0.727, 0.763] | 0.646 [0.628, 0.678] | 0.707 [0.678, 0.735] |
| Severity of 3 | 0.743 [0.698, 0.775] | 0.734 [0.696, 0.769] | 0.820 [0.793, 0.849] | 0.829 [0.797, 0.859] | **0.838 [0.812, 0.862]** | 0.801 [0.757, 0.836] | 0.727 [0.685, 0.761] | 0.753 [0.707, 0.791] |
| Averaged AUC | 0.713 [0.685, 0.740] | 0.698 [0.671, 0.723] | 0.774 [0.753, 0.797] | 0.773 [0.747, 0.795] | **0.791 [0.772, 0.812]** | 0.753 [0.728, 0.777] | 0.687 [0.660, 0.714] | 0.721 [0.692, 0.746] |

**Table 8. Pulmonary Edema Severity Results -** AUC across all severity levels for Finetuning and Linear Probing.

| FINETUNING | ImageNet-Pretrained Baseline | MedAug | CXR-RePaiR-CLIP | REFERS | ConVIRT | GLoRIA | S2MTS2 | MoCo-CXR |
|---|---|---|---|---|---|---|---|---|
| **Cardiomegaly** | 0.835 [0.804, 0.868] | 0.835 [0.805, 0.859] | 0.895 [0.863, 0.917] | 0.920 [0.898, 0.937] | **0.926 [0.904, 0.946]** | 0.873 [0.845, 0.900] | 0.885 [0.854, 0.906] | 0.846 [0.815, 0.874] |
| **Spondylosis** | 0.810 [0.711, 0.888] | 0.674 [0.597, 0.752] | 0.694 [0.614, 0.750] | **0.751 [0.682, 0.836]** | 0.706 [0.625, 0.780] | 0.710 [0.627, 0.781] | 0.749 [0.655, 0.862] | 0.682 [0.586, 0.761] |
| **Hernia and Hiatal** | 0.677 [0.545, 0.797] | 0.796 [0.725, 0.869] | 0.794 [0.714, 0.860] | **0.892 [0.833, 0.966]** | 0.837 [0.715, 0.956] | 0.743 [0.646, 0.851] | 0.810 [0.690, 0.892] | 0.741 [0.658, 0.819] |
| **Airspace Disease** | 0.807 [0.735, 0.860] | 0.800 [0.734, 0.847] | 0.798 [0.724, 0.857] | 0.844 [0.793, 0.892] | **0.877 [0.837, 0.910]** | 0.817 [0.748, 0.863] | 0.803 [0.733, 0.861] | 0.771 [0.692, 0.821] |
| **Pleural Effusion** | 0.859 [0.802, 0.908] | 0.911 [0.875, 0.948] | 0.911 [0.864, 0.944] | 0.928 [0.882, 0.965] | **0.946 [0.913, 0.975]** | 0.909 [0.875, 0.937] | 0.904 [0.867, 0.934] | 0.887 [0.840, 0.920] |
| **Costophrenic Angle** | 0.758 [0.657, 0.845] | 0.821 [0.742, 0.884] | 0.809 [0.727, 0.891] | 0.805 [0.730, 0.893] | **0.839 [0.772, 0.906]** | 0.819 [0.752, 0.891] | 0.756 [0.669, 0.839] | 0.766 [0.693, 0.837] |
| **Emphysema** | 0.831 [0.755, 0.897] | 0.862 [0.817, 0.912] | 0.843 [0.788, 0.903] | 0.894 [0.8386, 0.946] | **0.923 [0.880, 0.963]** | 0.848 [0.784, 0.903] | 0.904 [0.850, 0.947] | 0.854 [0.793, 0.900] |
| **Pulmonary Atelectasis** | 0.748 [0.710, 0.785] | 0.795 [0.759, 0.832] | 0.811 [0.774, 0.844] | 0.829 [0.793, 0.860] | **0.833 [0.797, 0.865]** | 0.790 [0.748, 0.836] | 0.787 [0.749, 0.838] | 0.792 [0.756, 0.838] |
| **Medical Device** | 0.715 [0.623, 0.804] | 0.680 [0.565, 0.783] | 0.699 [0.574, 0.800] | 0.762 [0.634, 0.884] | 0.761 [0.629, 0.855] | 0.660 [0.561, 0.740] | **0.770 [0.677, 0.856]** | 0.704 [0.608, 0.798] |
| **Scoliosis** | 0.644 [0.558, 0.720] | 0.602 [0.534, 0.670] | 0.531 [0.426, 0.601] | **0.744 [0.679, 0.802]** | 0.659 [0.589, 0.727] | 0.591 [0.509, 0.662] | 0.641 [0.551, 0.712] | 0.621 [0.546, 0.694] |
| **Nodule** | 0.571 [0.478, 0.650] | 0.536 [0.436, 0.609] | 0.513 [0.411, 0.589] | 0.615 [0.511, 0.687] | 0.601 [0.494, 0.694] | 0.539 [0.456, 0.608] | **0.632 [0.533, 0.702]** | 0.480 [0.383, 0.571] |
| **Atherosclerosis** | 0.771 [0.714, 0.840] | 0.788 [0.734, 0.842] | 0.751 [0.683, 0.809] | **0.830 [0.770, 0.872]** | 0.799 [0.733, 0.849] | 0.796 [0.742, 0.841] | 0.800 [0.746, 0.865] | 0.755 [0.683, 0.818] |
| **Calcinosis** | 0.650 [0.603, 0.710] | 0.627 [0.570, 0.681] | 0.607 [0.540, 0.662] | 0.682 [0.629, 0.734] | **0.686 [0.631, 0.739]** | 0.672 [0.604, 0.727] | 0.671 [0.609, 0.731] | 0.615 [0.557, 0.681] |
| **Pneumonia** | 0.561 [0.355, 0.776] | 0.627 [0.343, 0.832] | 0.630 [0.434, 0.814] | 0.763 [0.595, 0.899] | **0.769 [0.597, 0.911]** | 0.636 [0.396, 0.847] | 0.657 [0.329, 0.846] | 0.625 [0.344, 0.828] |
| **Granuloma** | 0.588 [0.544, 0.632] | 0.538 [0.498, 0.576] | 0.539 [0.489, 0.595] | **0.634 [0.594, 0.674]** | 0.627 [0.581, 0.673] | 0.570 [0.526, 0.618] | 0.602 [0.547, 0.640] | 0.525 [0.473, 0.576] |
| **Calcified Granuloma** | 0.605 [0.551, 0.662] | 0.519 [0.467, 0.558] | 0.531 [0.480, 0.592] | **0.634 [0.587, 0.699]** | 0.605 [0.550, 0.665] | 0.532 [0.473, 0.589] | 0.606 [0.551, 0.658] | 0.491 [0.437, 0.542] |
| **Pulmonary Congestion** | 0.816 [0.739, 0.884] | 0.826 [0.776, 0.888] | 0.834 [0.766, 0.906] | 0.894 [0.847, 0.943] | **0.897 [0.847, 0.948]** | 0.846 [0.782, 0.904] | 0.877 [0.823, 0.926] | 0.811 [0.740, 0.878] |
| **Opacity** | 0.731 [0.679, 0.769] | 0.778 [0.734, 0.812] | 0.784 [0.744, 0.816] | 0.812 [0.773, 0.851] | **0.819 [0.783, 0.854]** | 0.790 [0.753, 0.819] | 0.759 [0.718, 0.793] | 0.763 [0.716, 0.795] |
| **Pulmonary Edema** | 0.820 [0.732, 0.892] | 0.847 [0.791, 0.911] | 0.884 [0.803, 0.953] | 0.934 [0.861, 0.986] | **0.938 [0.906, 0.967]** | 0.857 [0.785, 0.923] | 0.936 [0.905, 0.964] | 0.848 [0.798, 0.901] |
| **normal** | 0.734 [0.705, 0.763] | 0.797 [0.771, 0.818] | 0.776 [0.748, 0.804] | 0.805 [0.781, 0.986] | **0.819 [0.797, 0.844]** | 0.786 [0.756, 0.811] | 0.789 [0.767, 0.809] | 0.773 [0.747, 0.798] |
| **Overall AUC** | 0.727 [0.650, 0.797] | 0.733 [0.664, 0.794] | 0.732 [0.658, 0.795] | **0.799 [0.736, 0.866]** | 0.793 [0.729, 0.851] | 0.739 [0.668, 0.803] | 0.767 [0.691, 0.829] | 0.718 [0.643, 0.782] |

| LINEAR PROBING | ImageNet-Pretrained Baseline | MedAug | CXR-RePaiR-CLIP | REFERS | ConVIRT | GLoRIA | S2MTS2 | MoCo-CXR |
|---|---|---|---|---|---|---|---|---|
| **Cardiomegaly** | 0.835 [0.804, 0.868] | 0.850 [0.809, 0.881] | 0.904 [0.875, 0.927] | 0.921 [0.893, 0.939] | **0.923 [0.896, 0.946]** | 0.881 [0.844, 0.908] | 0.862 [0.826, 0.887] | 0.854 [0.825, 0.882] |
| **Spondylosis** | 0.810 [0.711, 0.888] | 0.666 [0.586, 0.757] | 0.744 [0.657, 0.852] | 0.706 [0.596, 0.814] | 0.711 [0.605, 0.824] | **0.773 [0.682, 0.875]** | 0.686 [0.590, 0.789] | 0.758 [0.674, 0.850] |
| **Hernia and Hiatal** | 0.677 [0.545, 0.797] | 0.850 [0.746, 0.936] | 0.860 [0.783, 0.926] | **0.907 [0.858, 0.963]** | 0.930 [0.840, 0.993] | 0.755 [0.652, 0.865] | 0.784 [0.636, 0.889] | 0.813 [0.749, 0.881] |
| **Airspace Disease** | 0.807 [0.735, 0.860] | 0.797 [0.729, 0.854] | 0.849 [0.778, 0.896] | 0.872 [0.821, 0.914] | **0.900 [0.863, 0.928]** | 0.796 [0.736, 0.855] | 0.777 [0.695, 0.835] | 0.738 [0.642, 0.815] |
| **Pleural Effusion** | 0.859 [0.802, 0.908] | 0.912 [0.877, 0.943] | 0.930 [0.893, 0.957] | 0.931 [0.900, 0.956] | **0.950 [0.916, 0.976]** | 0.880 [0.839, 0.908] | 0.885 [0.846, 0.927] | 0.865 [0.824, 0.900] |
| **Costophrenic Angle** | 0.758 [0.657, 0.845] | 0.840 [0.792, 0.890] | 0.830 [0.766, 0.896] | **0.831 [0.772, 0.894]** | 0.826 [0.746, 0.893] | 0.767 [0.683, 0.847] | 0.737 [0.670, 0.815] | 0.671 [0.597, 0.762] |
| **Emphysema** | 0.831 [0.755, 0.897] | 0.839 [0.775, 0.905] | 0.881 [0.807, 0.945] | 0.863 [0.778, 0.937] | **0.897 [0.808, 0.969]** | 0.841 [0.757, 0.917] | 0.853 [0.785, 0.920] | 0.840 [0.743, 0.912] |
| **Pulmonary Atelectasis** | 0.748 [0.710, 0.785] | 0.806 [0.766, 0.840] | 0.832 [0.790, 0.867] | 0.832 [0.796, 0.863] | **0.845 [0.806, 0.874]** | 0.767 [0.723, 0.822] | 0.783 [0.738, 0.830] | 0.743 [0.706, 0.782] |
| **Medical Device** | 0.715 [0.623, 0.804] | 0.731 [0.595, 0.828] | 0.756 [0.6237, 0.854] | **0.804 [0.691, 0.890]** | 0.798 [0.655, 0.889] | 0.736 [0.615, 0.835] | **0.770 [0.679, 0.867]** | 0.772 [0.688, 0.869] |
| **Scoliosis** | 0.644 [0.558, 0.720] | 0.645 [0.574, 0.708] | 0.569 [0.485, 0.631] | **0.806 [0.759, 0.855]** | 0.707 [0.626, 0.792] | 0.662 [0.599, 0.731] | 0.610 [0.499, 0.695] | 0.632 [0.539, 0.712] |
| **Nodule** | 0.571 [0.478, 0.650] | 0.646 [0.548, 0.710] | 0.665 [0.587, 0.718] | 0.637 [0.558, 0.713] | **0.657 [0.560, 0.748]** | 0.659 [0.562, 0.756] | **0.587 [0.511, 0.661]** | 0.597 [0.521, 0.677] |
| **Atherosclerosis** | 0.771 [0.714, 0.840] | 0.781 [0.709, 0.845] | 0.813 [0.750, 0.861] | 0.831 [0.771, 0.871] | **0.855 [0.812, 0.891]** | 0.788 [0.713, 0.844] | 0.763 [0.698, 0.828] | 0.730 [0.650, 0.798] |
| **Calcinosis** | 0.650 [0.603, 0.710] | 0.651 [0.588, 0.715] | 0.676 [0.629, 0.726] | 0.703 [0.651, 0.750] | **0.715 [0.665, 0.760]** | 0.652 [0.585, 0.720] | 0.654 [0.600, 0.710] | 0.626 [0.573, 0.688] |
| **Pneumonia** | 0.561 [0.355, 0.776] | 0.654 [0.454, 0.808] | 0.731 [0.532, 0.864] | 0.806 [0.701, 0.910] | **0.878 [0.725, 0.967]** | 0.605 [0.395, 0.844] | 0.598 [0.333, 0.807] | 0.590 [0.363, 0.800] |
| **Granuloma** | 0.588 [0.544, 0.632] | 0.622 [0.559, 0.650] | 0.600 [0.550, 0.646] | 0.637 [0.594, 0.672] | **0.639 [0.594, 0.689]** | 0.577 [0.532, 0.622] | 0.591 [0.544, 0.631] | 0.590 [0.550, 0.637] |
| **Calcified Granuloma** | 0.605 [0.551, 0.662] | 0.600 [0.554, 0.656] | 0.606 [0.550, 0.661] | 0.631 [0.583, 0.679] | **0.633 [0.583, 0.694]** | 0.544 [0.482, 0.603] | 0.590 [0.534, 0.650] | 0.552 [0.508, 0.613] |
| **Pulmonary Congestion** | 0.816 [0.739, 0.884] | 0.857 [0.800, 0.912] | 0.898 [0.856, 0.942] | 0.905 [0.861, 0.953] | **0.932 [0.904, 0.962]** | 0.868 [0.811, 0.936] | 0.856 [0.796, 0.915] | 0.834 [0.768, 0.906] |
| **Opacity** | 0.731 [0.679, 0.769] | 0.766 [0.720, 0.801] | 0.788 [0.751, 0.817] | 0.812 [0.772, 0.846] | **0.837 [0.808, 0.865]** | 0.787 [0.755, 0.816] | 0.730 [0.678, 0.767] | 0.725 [0.680, 0.762] |
| **Pulmonary Edema** | 0.820 [0.732, 0.892] | 0.888 [0.853, 0.921] | 0.943 [0.897, 0.982] | 0.950 [0.902, 0.986] | **0.979 [0.971, 0.991]** | 0.898 [0.803, 0.982] | 0.936 [0.894, 0.980] | 0.877 [0.805, 0.942] |
| **normal** | 0.734 [0.705, 0.763] | 0.778 [0.749, 0.798] | 0.778 [0.752, 0.804] | 0.803 [0.776, 0.830] | **0.820 [0.793, 0.844]** | 0.778 [0.756, 0.803] | 0.752 [0.727, 0.778] | 0.744 [0.718, 0.771] |
| **Overall AUC** | 0.727 [0.650, 0.797] | 0.759 [0.689, 0.818] | 0.783 [0.716, 0.839] | 0.809 [0.752, 0.862] | **0.822 [0.759, 0.875]** | 0.751 [0.676, 0.824] | 0.740 [0.664, 0.809] | 0.728 [0.656, 0.798] |

**Table 9. Indiana Results -** AUC across all diagnoses for Finetuning and Linear Probing.

| FINETUNING | ImageNet-Pretrained Baseline | MedAug | CXR-RePaiR-CLIP | REFERS | ConVIRT | GLoRIA | S2MTS2 | MoCo-CXR |
|---|---|---|---|---|---|---|---|---|
| Subtlety of 0 | 0.536 [0.397, 0.642] | 0.688 [0.546, 0.772] | 0.704 [0.582, 0.813] | 0.739 [0.650, 0.833] | 0.673 [0.554, 0.755] | **0.757 [0.656, 0.856]** | 0.740 [0.650, 0.819] | **0.757 [0.659, 0.845]** |
| Subtlety of 1 | 0.452 [0.276, 0.629] | 0.470 [0.307, 0.658] | 0.607 [0.446, 0.783] | 0.616 [0.415, 0.773] | 0.467 [0.329, 0.636] | **0.637 [0.529, 0.797]** | 0.426 [0.285, 0.582] | 0.610 [0.473, 0.756] |
| Subtlety of 2 | 0.566 [0.361, 0.719] | 0.560 [0.405, 0.733] | 0.595 [0.435, 0.739] | 0.625 [0.507, 0.738] | **0.628 [0.542, 0.793]** | 0.554 [0.395, 0.724] | 0.583 [0.415, 0.705] | 0.470 [0.310, 0.694] |
| Subtlety of 3 | 0.445 [0.275, 0.659] | 0.397 [0.286, 0.588] | 0.396 [0.276, 0.550] | 0.478 [0.302, 0.658] | **0.500 [0.358, 0.601]** | 0.311 [0.200, 0.428] | 0.427 [0.324, 0.565] | 0.491 [0.317, 0.627] |
| Subtlety of 4 | 0.544 [0.305, 0.729] | 0.483 [0.359, 0.612] | 0.388 [0.272, 0.511] | 0.364 [0.160, 0.524] | **0.531 [0.340, 0.705]** | 0.381 [0.267, 0.487] | 0.490 [0.370, 0.597] | 0.371 [0.242, 0.496] |
| Subtlety of 5 | **0.716 [0.465, 0.879]** | 0.657 [0.305, 0.927] | 0.378 [0.159, 0.534] | 0.431 [0.164, 0.700] | 0.529 [0.422, 0.625] | 0.681 [0.454, 0.881] | 0.270 [0.055, 0.545] | 0.652 [0.330, 0.867] |
| Overall AUC | 0.543 [0.347, 0.709] | 0.542 [0.368, 0.715] | 0.511 [0.362, 0.655] | 0.542 [0.366, 0.704] | 0.555 [0.424, 0.686] | 0.554 [0.417, 0.696] | 0.489 [0.350, 0.635] | **0.558 [0.388, 0.714]** |

| LINEAR PROBING | ImageNet-Pretrained Baseline | MedAug | CXR-RePaiR-CLIP | REFERS | ConVIRT | GLoRIA | S2MTS2 | MoCo-CXR |
|---|---|---|---|---|---|---|---|---|
| Subtlety of 0 | 0.536 [0.397, 0.642] | 0.607 [0.494, 0.727] | **0.698 [0.572, 0.805]** | 0.622 [0.505, 0.728] | 0.590 [0.466, 0.696] | 0.577 [0.466, 0.694] | 0.553 [0.450, 0.671] | 0.623 [0.500, 0.720] |
| Subtlety of 1 | 0.452 [0.276, 0.629] | **0.679 [0.541, 0.839]** | 0.509 [0.370, 0.676] | 0.569 [0.414, 0.770] | 0.563 [0.418, 0.711] | 0.628 [0.477, 0.752] | 0.369 [0.227, 0.542] | 0.539 [0.390, 0.724] |
| Subtlety of 2 | 0.566 [0.361, 0.719] | **0.735 [0.617, 0.865]** | 0.595 [0.442, 0.764] | 0.688 [0.573, 0.787] | 0.661 [0.552, 0.824] | 0.586 [0.411, 0.786] | 0.563 [0.416, 0.709] | 0.476 [0.319, 0.655] |
| Subtlety of 3 | 0.445 [0.275, 0.659] | 0.451 [0.322, 0.592] | 0.462 [0.354, 0.619] | **0.526 [0.400, 0.653]** | 0.368 [0.233, 0.484] | 0.518 [0.381, 0.673] | 0.388 [0.235, 0.501] | 0.447 [0.350, 0.555] |
| Subtlety of 4 | 0.544 [0.305, 0.729] | 0.459 [0.254, 0.649] | 0.446 [0.317, 0.546] | 0.490 [0.313, 0.661] | 0.367 [0.209, 0.527] | 0.456 [0.227, 0.666] | 0.483 [0.338, 0.589] | 0.463 [0.290, 0.615] |
| Subtlety of 5 | **0.716 [0.465, 0.879]** | 0.520 [0.319, 0.714] | 0.422 [0.154, 0.680] | 0.412 [0.165, 0.632] | 0.574 [0.408, 0.773] | **0.755 [0.552, 0.912]** | 0.309 [0.069, 0.557] | 0.613 [0.293, 0.838] |
| Overall AUC | 0.543 [0.347, 0.709] | 0.575 [0.425, 0.731] | 0.522 [0.368, 0.682] | 0.551 [0.395, 0.705] | 0.520 [0.381, 0.669] | **0.587 [0.419, 0.747]** | 0.444 [0.289, 0.595] | 0.527 [0.357, 0.685] |

**Table 10. JSRT Results -** AUC across all subtleties for Finetuning and Linear Probing.

| FINETUNING | ImageNet-Pretrained Baseline | MedAug | CXR-RePaiR-CLIP | REFERS | ConVIRT | GLoRIA | S2MTS2 | MoCo-CXR |
|---|---|---|---|---|---|---|---|---|
| Atelectasis | 0.774 [0.726, 0.805] | 0.775 [0.740, 0.806] | 0.817 [0.772, 0.852] | 0.833 [0.789, 0.859] | **0.844 [0.810, 0.873]** | 0.840 [0.803, 0.875] | 0.790 [0.742, 0.833] | 0.751 [0.715, 0.781] |
| Consolidation | 0.785 [0.747, 0.813] | 0.796 [0.758, 0.832] | 0.851 [0.819, 0.878] | 0.851 [0.809, 0.872] | **0.861 [0.825, 0.893]** | 0.844 [0.804, 0.876] | 0.801 [0.764, 0.837] | 0.805 [0.764, 0.837] |
| Enlarged cardiac silhouette | 0.809 [0.777, 0.847] | 0.790 [0.754, 0.828] | 0.884 [0.853, 0.913] | 0.900 [0.883, 0.941] | **0.922 [0.903, 0.946]** | 0.907 [0.876, 0.937] | 0.836 [0.799, 0.872] | 0.817 [0.789, 0.853] |
| Groundglass opacity | 0.693 [0.619, 0.743] | 0.683 [0.627, 0.753] | 0.732 [0.677, 0.781] | 0.735 [0.665, 0.784] | **0.752 [0.691, 0.799]** | 0.750 [0.691, 0.796] | 0.697 [0.634, 0.758] | 0.699 [0.637, 0.756] |
| High lung volume / emphysema | 0.772 [0.676, 0.875] | 0.798 [0.670, 0.903] | 0.841 [0.748, 0.913] | 0.899 [0.859, 0.950] | 0.858 [0.781, 0.948] | 0.857 [0.787, 0.926] | **0.918 [0.801, 0.982]** | 0.692 [0.577, 0.797] |
| Lung nodule or mass | 0.630 [0.513, 0.745] | 0.554 [0.445, 0.668] | 0.668 [0.556, 0.769] | 0.688 [0.582, 0.808] | 0.728 [0.635, 0.842] | **0.732 [0.627, 0.828]** | 0.647 [0.561, 0.774] | 0.632 [0.541, 0.738] |
| Pleural abnormality | 0.831 [0.793, 0.864] | 0.850 [0.818, 0.882] | 0.880 [0.850, 0.909] | 0.908 [0.884, 0.930] | **0.925 [0.897, 0.946]** | **0.925 [0.899, 0.944]** | 0.846 [0.818, 0.878] | 0.830 [0.803, 0.867] |
| Pneumothorax | 0.584 [0.442, 0.688] | 0.702 [0.583, 0.792] | 0.767 [0.634, 0.896] | 0.770 [0.696, 0.910] | 0.839 [0.733, 0.963] | **0.840 [0.740, 0.937]** | 0.739 [0.618, 0.857] | 0.665 [0.522, 0.775] |
| Pulmonary edema | 0.817 [0.776, 0.853] | 0.779 [0.736, 0.828] | 0.859 [0.812, 0.892] | 0.844 [0.829, 0.900] | 0.892 [0.858, 0.920] | **0.896 [0.864, 0.923]** | 0.856 [0.806, 0.901] | 0.801 [0.756, 0.844] |
| Overall AUC | 0.744 [0.674, 0.804] | 0.747 [0.681, 0.810] | 0.811 [0.747, 0.867] | 0.825 [0.777, 0.884] | **0.847 [0.793, 0.903]** | 0.843 [0.788, 0.894] | 0.792 [0.727, 0.855] | 0.744 [0.678, 0.805] |

| LINEAR PROBING | ImageNet-Pretrained Baseline | MedAug | CXR-RePaiR-CLIP | REFERS | ConVIRT | GLoRIA | S2MTS2 | MoCo-CXR |
|---|---|---|---|---|---|---|---|---|
| Atelectasis | 0.774 [0.726, 0.805] | 0.769 [0.724, 0.804] | 0.830 [0.790, 0.863] | 0.834 [0.797, 0.863] | **0.849 [0.808, 0.879]** | 0.845 [0.803, 0.879] | 0.786 [0.746, 0.822] | 0.744 [0.701, 0.785] |
| Consolidation | 0.785 [0.747, 0.813] | 0.774 [0.730, 0.819] | 0.855 [0.818, 0.885] | 0.855 [0.820, 0.883] | **0.861 [0.819, 0.888]** | 0.856 [0.816, 0.888] | 0.782 [0.742, 0.824] | 0.782 [0.735, 0.819] |
| Enlarged cardiac silhouette | 0.809 [0.777, 0.847] | 0.814 [0.776, 0.847] | 0.898 [0.873, 0.920] | 0.904 [0.881, 0.937] | **0.918 [0.896, 0.938]** | 0.907 [0.8788, 0.934] | 0.801 [0.771, 0.836] | 0.828 [0.795, 0.861] |
| Groundglass opacity | 0.693 [0.619, 0.743] | 0.687 [0.617, 0.744] | 0.744 [0.676, 0.808] | 0.742 [0.694, 0.794] | **0.754 [0.693, 0.800]** | 0.753 [0.699, 0.808] | 0.689 [0.638, 0.747] | 0.701 [0.645, 0.761] |
| High lung volume / emphysema | 0.772 [0.676, 0.875] | 0.883 [0.788, 0.959] | 0.872 [0.765, 0.963] | **0.950 [0.903, 0.982]** | 0.916 [0.842, 0.971] | 0.919 [0.868, 0.964] | 0.919 [0.812, 0.991] | 0.865 [0.787, 0.934] |
| Lung nodule or mass | 0.630 [0.513, 0.745] | 0.603 [0.507, 0.698] | 0.666 [0.571, 0.785] | 0.739 [0.651, 0.825] | 0.733 [0.618, 0.821] | **0.784 [0.710, 0.862]** | 0.651 [0.556, 0.788] | 0.644 [0.536, 0.761] |
| Pleural abnormality | 0.831 [0.793, 0.864] | 0.855 [0.826, 0.889] | 0.905 [0.877, 0.927] | 0.907 [0.878, 0.927] | 0.919 [0.893, 0.941] | **0.925 [0.899, 0.943]** | 0.812 [0.782, 0.849] | 0.810 [0.773, 0.846] |
| Pneumothorax | 0.584 [0.442, 0.688] | 0.766 [0.656, 0.854] | 0.835 [0.725, 0.943] | 0.809 [0.678, 0.936] | 0.772 [0.633, 0.942] | **0.799 [0.671, 0.928]** | 0.684 [0.556, 0.801] | 0.549 [0.440, 0.688] |
| Pulmonary edema | 0.817 [0.776, 0.853] | 0.806 [0.755, 0.845] | 0.878 [0.842, 0.910] | 0.867 [0.828, 0.900] | **0.897 [0.862, 0.924]** | 0.888 [0.854, 0.918] | 0.838 [0.798, 0.877] | 0.828 [0.776, 0.862] |
| Overall AUC | 0.744 [0.674, 0.804] | 0.773 [0.709, 0.829] | 0.831 [0.771, 0.889] | 0.845 [0.792, 0.894] | 0.847 [0.785, 0.900] | **0.853 [0.800, 0.903]** | 0.774 [0.711, 0.837] | 0.750 [0.688, 0.813] |

**Table 11. REFLACX Results -** AUC across all diagnoses for Finetuning and Linear Probing.

| FINETUNING | ImageNet-Pretrained Baseline | MedAug | CXR-RePaiR-CLIP | REFERS | ConVIRT | GLoRIA | S2MTS2 | MoCo-CXR |
|---|---|---|---|---|---|---|---|---|
| No Finding | 0.762 [0.753, 0.773] | 0.776 [0.766, 0.786] | 0.783 [0.774, 0.794] | 0.800 [0.790, 0.810] | **0.804 [0.795, 0.814]** | 0.786 [0.776, 0.796] | 0.784 [0.774, 0.795] | 0.775 [0.766, 0.784] |
| Enlarged Cardiomediastinum | 0.735 [0.612, 0.845] | 0.751 [0.650, 0.862] | 0.753 [0.613, 0.863] | 0.740 [0.647, 0.840] | **0.767 [0.681, 0.877]** | 0.681 [0.575, 0.827] | 0.760 [0.659, 0.889] | 0.726 [0.621, 0.840] |
| Cardiomegaly | 0.855 [0.843, 0.867] | 0.871 [0.860, 0.883] | 0.902 [0.892, 0.914] | 0.917 [0.908, 0.926] | **0.918 [0.910, 0.928]** | 0.899 [0.890, 0.909] | 0.902 [0.892, 0.912] | 0.876 [0.866, 0.888] |
| Lung Lesion | 0.563 [0.540, 0.601] | 0.593 [0.564, 0.634] | 0.586 [0.559, 0.622] | 0.624 [0.595, 0.655] | **0.631 [0.601, 0.667]** | 0.603 [0.574, 0.633] | 0.597 [0.564, 0.627] | 0.578 [0.542, 0.613] |
| Lung Opacity | 0.732 [0.716, 0.751] | 0.748 [0.730, 0.762] | 0.750 [0.730, 0.771] | 0.786 [0.772, 0.807] | **0.792 [0.774, 0.807]** | 0.757 [0.738, 0.773] | 0.744 [0.729, 0.762] | 0.737 [0.721, 0.758] |
| Edema | 0.782 [0.567, 0.952] | 0.747 [0.548, 0.909] | 0.750 [0.547, 0.929] | 0.762 [0.584, 0.917] | 0.757 [0.579, 0.918] | 0.782 [0.641, 0.889] | **0.812 [0.611, 0.954]** | 0.751 [0.560, 0.894] |
| Consolidation | 0.801 [0.780, 0.820] | 0.795 [0.779, 0.816] | 0.803 [0.785, 0.825] | **0.816 [0.793, 0.830]** | 0.813 [0.794, 0.831] | 0.812 [0.793, 0.829] | 0.796 [0.777, 0.813] | 0.795 [0.773, 0.815] |
| Pneumonia | 0.819 [0.788, 0.850] | 0.847 [0.822, 0.870] | 0.847 [0.819, 0.876] | **0.899 [0.877, 0.922]** | 0.885 [0.860, 0.908] | 0.850 [0.816, 0.877] | 0.848 [0.822, 0.875] | 0.835 [0.804, 0.861] |
| Atelectasis | 0.773 [0.759, 0.794] | 0.782 [0.770, 0.802] | 0.796 [0.786, 0.814] | 0.823 [0.810, 0.840] | **0.843 [0.830, 0.859]** | 0.808 [0.797, 0.828] | 0.802 [0.791, 0.821] | 0.778 [0.766, 0.797] |
| Pneumothorax | 0.885 [0.820, 0.937] | 0.904 [0.864, 0.939] | 0.850 [0.751, 0.919] | 0.801 [0.716, 0.883] | **0.926 [0.886, 0.964]** | 0.859 [0.782, 0.937] | 0.865 [0.785, 0.943] | 0.831 [0.776, 0.887] |
| Pleural Effusion | 0.863 [0.847, 0.881] | 0.878 [0.864, 0.892] | 0.899 [0.881, 0.913] | 0.917 [0.903, 0.933] | **0.929 [0.916, 0.940]** | 0.899 [0.885, 0.913] | 0.883 [0.868, 0.897] | 0.866 [0.851, 0.881] |
| Pleural Other | **0.889 [0.851, 0.931]** | 0.830 [0.760, 0.896] | 0.877 [0.820, 0.927] | 0.867 [0.809, 0.927] | 0.842 [0.766, 0.900] | 0.841 [0.756, 0.895] | 0.881 [0.821, 0.929] | 0.866 [0.815, 0.912] |
| Fracture | 0.623 [0.582, 0.660] | 0.648 [0.613, 0.699] | 0.650 [0.609, 0.704] | 0.669 [0.633, 0.713] | **0.683 [0.640, 0.733]** | 0.680 [0.636, 0.724] | 0.683 [0.645, 0.725] | 0.634 [0.591, 0.678] |
| Support Devices | 0.827 [0.817, 0.836] | 0.855 [0.846, 0.867] | 0.846 [0.836, 0.857] | **0.892 [0.887, 0.901]** | 0.892 [0.883, 0.903] | 0.881 [0.871, 0.891] | 0.864 [0.857, 0.874] | 0.859 [0.851, 0.871] |
| Overall AUC | 0.779 [0.734, 0.821] | 0.787 [0.745, 0.830] | 0.792 [0.743, 0.838] | 0.808 [0.766, 0.850] | **0.820 [0.780, 0.861]** | 0.796 [0.752, 0.837] | 0.801 [0.757, 0.844] | 0.779 [0.736, 0.820] |

| LINEAR PROBING | ImageNet-Pretrained Baseline | MedAug | CXR-RePaiR-CLIP | REFERS | ConVIRT | GLoRIA | S2MTS2 | MoCo-CXR |
|---|---|---|---|---|---|---|---|---|
| No Finding | 0.762 [0.753, 0.773] | 0.749 [0.739, 0.760] | 0.765 [0.756, 0.774] | 0.785 [0.776, 0.797] | **0.798 [0.789, 0.808]** | 0.781 [0.771, 0.790] | 0.745 [0.734, 0.757] | 0.741 [0.730, 0.752] |
| Enlarged Cardiomediastinum | 0.735 [0.612, 0.845] | 0.699 [0.594, 0.831] | 0.724 [0.588, 0.859] | 0.681 [0.567, 0.836] | 0.679 [0.568, 0.828] | **0.761 [0.659, 0.855]** | 0.710 [0.605, 0.838] | 0.696 [0.590, 0.814] |
| Cardiomegaly | 0.855 [0.843, 0.867] | 0.863 [0.853, 0.875] | 0.892 [0.878, 0.903] | 0.910 [0.898, 0.922] | **0.916 [0.907, 0.925]** | 0.892 [0.880, 0.903] | 0.868 [0.856, 0.882] | 0.852 [0.839, 0.865] |
| Lung Lesion | 0.563 [0.540, 0.601] | 0.606 [0.581, 0.638] | 0.626 [0.601, 0.656] | 0.607 [0.575, 0.637] | 0.644 [0.619, 0.681] | **0.656 [0.632, 0.687]** | 0.597 [0.569, 0.625] | 0.624 [0.594, 0.657] |
| Lung Opacity | 0.732 [0.716, 0.751] | 0.721 [0.705, 0.740] | 0.748 [0.734, 0.766] | 0.774 [0.759, 0.793] | **0.786 [0.769, 0.805]** | 0.760 [0.745, 0.777] | 0.706 [0.690, 0.725] | 0.712 [0.696, 0.732] |
| Edema | 0.782 [0.567, 0.952] | 0.771 [0.576, 0.947] | **0.832 [0.675, 0.930]** | 0.785 [0.675, 0.901] | 0.806 [0.643, 0.936] | 0.764 [0.623, 0.876] | 0.749 [0.545, 0.943] | 0.741 [0.551, 0.946] |
| Consolidation | 0.801 [0.780, 0.820] | 0.764 [0.743, 0.783] | 0.794 [0.778, 0.814] | 0.799 [0.774, 0.818] | **0.807 [0.787, 0.826]** | 0.796 [0.779, 0.816] | 0.764 [0.746, 0.788] | 0.785 [0.760, 0.804] |
| Pneumonia | 0.819 [0.788, 0.850] | 0.786 [0.753, 0.822] | 0.836 [0.807, 0.860] | 0.872 [0.850, 0.895] | **0.884 [0.864, 0.909]** | 0.832 [0.801, 0.864] | 0.809 [0.779, 0.837] | 0.798 [0.760, 0.830] |
| Atelectasis | 0.773 [0.759, 0.794] | 0.767 [0.757, 0.789] | 0.792 [0.781, 0.814] | 0.812 [0.798, 0.827] | **0.829 [0.817, 0.847]** | 0.799 [0.786, 0.816] | 0.752 [0.738, 0.773] | 0.747 [0.731, 0.766] |
| Pneumothorax | 0.885 [0.820, 0.937] | 0.853 [0.789, 0.916] | 0.897 [0.830, 0.959] | 0.853 [0.776, 0.941] | **0.942 [0.894, 0.977]** | 0.882 [0.801, 0.963] | 0.863 [0.784, 0.948] | 0.909 [0.842, 0.971] |
| Pleural Effusion | 0.863 [0.847, 0.881] | 0.890 [0.873, 0.904] | 0.901 [0.885, 0.916] | 0.912 [0.898, 0.924] | **0.930 [0.918, 0.943]** | 0.897 [0.880, 0.913] | 0.838 [0.817, 0.859] | 0.863 [0.842, 0.885] |
| Pleural Other | 0.889 [0.851, 0.931] | 0.887 [0.850, 0.928] | 0.897 [0.857, 0.932] | 0.895 [0.867, 0.923] | **0.913 [0.878, 0.937]** | 0.753 [0.604, 0.825] | 0.864 [0.812, 0.905] | 0.790 [0.699, 0.882] |
| Fracture | 0.623 [0.582, 0.660] | 0.668 [0.636, 0.700] | 0.677 [0.639, 0.727] | **0.697 [0.648, 0.745]** | 0.689 [0.650, 0.729] | 0.671 [0.631, 0.715] | 0.659 [0.611, 0.708] | 0.642 [0.604, 0.680] |
| Support Devices | 0.827 [0.817, 0.836] | 0.841 [0.832, 0.853] | 0.812 [0.804, 0.826] | 0.864 [0.854, 0.875] | **0.868 [0.860, 0.880]** | 0.862 [0.853, 0.874] | 0.807 [0.795, 0.820] | 0.807 [0.798, 0.820] |
| Overall AUC | 0.779 [0.734, 0.821] | 0.776 [0.734, 0.820] | 0.799 [0.758, 0.838] | 0.803 [0.765, 0.845] | **0.821 [0.783, 0.859]** | 0.793 [0.746, 0.834] | 0.767 [0.720, 0.815] | 0.765 [0.717, 0.815] |

**Table 12. BRAX Results -** AUC across all diagnoses for Finetuning and Linear Probing.

| FINETUNING | ImageNet-Pretrained Baseline | MedAug | CXR-RePaiR-CLIP | REFERS | ConVIRT | GLoRIA | S2MTS2 | MoCo-CXR |
|---|---|---|---|---|---|---|---|---|
| **Atelectasis** | 0.635 [0.616, 0.662] | 0.618 [0.599, 0.644] | 0.858 [0.844, 0.873] | 0.885 [0.868, 0.900] | **0.886 [0.872, 0.900]** | 0.884 [0.864, 0.900] | 0.819 [0.803, 0.838] | 0.630 [0.607, 0.653] |
| **Cardiomegaly** | 0.770 [0.749, 0.788] | 0.752 [0.732, 0.773] | 0.837 [0.819, 0.856] | 0.903 [0.889, 0.919] | **0.925 [0.912, 0.936]** | 0.919 [0.899, 0.931] | 0.848 [0.823, 0.861] | 0.747 [0.728, 0.764] |
| **Effusion** | 0.832 [0.825, 0.840] | 0.867 [0.861, 0.875] | 0.878 [0.871, 0.884] | 0.891 [0.885, 0.897] | **0.899 [0.893, 0.904]** | 0.897 [0.890, 0.903] | 0.857 [0.849, 0.865] | 0.864 [0.858, 0.871] |
| **Infiltration** | 0.628 [0.539, 0.733] | 0.612 [0.511, 0.742] | 0.624 [0.514, 0.746] | 0.711 [0.616, 0.834] | 0.742 [0.654, 0.834] | 0.638 [0.532, 0.765] | **0.752 [0.643, 0.829]** | 0.607 [0.507, 0.720] |
| **Mass** | 0.602 [0.593, 0.610] | 0.605 [0.596, 0.614] | 0.619 [0.608, 0.630] | **0.633 [0.624, 0.641]** | 0.627 [0.618, 0.636] | 0.633 [0.622, 0.640] | 0.624 [0.614, 0.634] | 0.608 [0.598, 0.617] |
| **Nodule** | 0.687 [0.672, 0.703] | 0.711 [0.696, 0.727] | 0.775 [0.759, 0.789] | 0.849 [0.833, 0.860] | **0.865 [0.854, 0.878]** | 0.844 [0.828, 0.854] | 0.748 [0.730, 0.768] | 0.704 [0.690, 0.721] |
| **Pneumonia** | 0.617 [0.602, 0.636] | 0.645 [0.631, 0.663] | 0.665 [0.650, 0.681] | 0.710 [0.694, 0.727] | **0.736 [0.722, 0.750]** | 0.720 [0.703, 0.740] | 0.664 [0.650, 0.681] | 0.647 [0.632, 0.669] |
| **Pneumothorax** | 0.740 [0.730, 0.752] | 0.752 [0.741, 0.761] | 0.789 [0.780, 0.799] | 0.804 [0.797, 0.812] | **0.821 [0.813, 0.827]** | 0.806 [0.799, 0.815] | 0.779 [0.747, 0.781] | 0.750 [0.740, 0.760] |
| **Consolidation** | 0.782 [0.771, 0.797] | 0.781 [0.768, 0.791] | 0.858 [0.847, 0.868] | 0.870 [0.858, 0.879] | **0.894 [0.883, 0.902]** | 0.886 [0.875, 0.896] | 0.854 [0.842, 0.868] | 0.775 [0.763, 0.787] |
| **Edema** | 0.722 [0.707, 0.736] | 0.743 [0.726, 0.759] | 0.771 [0.758, 0.794] | 0.781 [0.764, 0.798] | 0.786 [0.768, 0.802] | **0.787 [0.771, 0.802]** | 0.763 [0.747, 0.781] | 0.707 [0.687, 0.727] |
| **Emphysema** | 0.690 [0.661, 0.713] | 0.704 [0.672, 0.741] | 0.702 [0.675, 0.742] | 0.749 [0.712, 0.779] | **0.770 [0.733, 0.797]** | 0.744 [0.704, 0.770] | 0.717 [0.680, 0.745] | 0.699 [0.668, 0.729] |
| **Fibrosis** | 0.628 [0.585, 0.675] | 0.655 [0.603, 0.694] | 0.677 [0.642, 0.725] | 0.692 [0.647, 0.738] | **0.731 [0.699, 0.767]** | 0.688 [0.565, 0.736] | 0.713 [0.676, 0.742] | 0.599 [0.549, 0.651] |
| **Pleural Thickening** | 0.803 [0.784, 0.823] | 0.814 [0.794, 0.829] | 0.867 [0.850, 0.884] | 0.903 [0.888, 0.915] | **0.910 [0.897, 0.922]** | 0.898 [0.881, 0.910] | 0.879 [0.861, 0.892] | 0.795 [0.775, 0.815] |
| **Hernia** | 0.748 [0.734, 0.771] | 0.765 [0.754, 0.783] | 0.773 [0.759, 0.789] | 0.788 [0.774, 0.805] | **0.796 [0.783, 0.813]** | 0.785 [0.771, 0.804] | 0.773 [0.760, 0.793] | 0.761 [0.748, 0.779] |
| **Overall AUC** | 0.706 [0.683, 0.731] | 0.716 [0.692, 0.743] | 0.764 [0.741, 0.790] | 0.798 [0.775, 0.822] | **0.813 [0.793, 0.833]** | 0.795 [0.765, 0.819] | 0.771 [0.745, 0.791] | 0.707 [0.682, 0.733] |

| LINEAR PROBING | ImageNet-Pretrained Baseline | MedAug | CXR-RePaiR-CLIP | REFERS | ConVIRT | GLoRIA | S2MTS2 | MoCo-CXR |
|---|---|---|---|---|---|---|---|---|
| **Atelectasis** | 0.635 [0.616, 0.662] | 0.512 [0.480, 0.544] | **0.879 [0.865, 0.892]** | 0.876 [0.858, 0.895] | 0.819 [0.802, 0.841] | 0.756 [0.736, 0.783] | 0.539 [0.516, 0.570] | 0.600 [0.575, 0.628] |
| **Cardiomegaly** | 0.770 [0.749, 0.788] | 0.612 [0.587, 0.638] | 0.838 [0.819, 0.854] | 0.890 [0.871, 0.907] | 0.840 [0.822, 0.857] | **0.899 [0.881, 0.916]** | 0.533 [0.507, 0.563] | 0.646 [0.621, 0.673] |
| **Effusion** | 0.832 [0.825, 0.840] | 0.640 [0.628, 0.651] | 0.881 [0.874, 0.887] | **0.890 [0.883, 0.897]** | 0.719 [0.705, 0.732] | 0.706 [0.694, 0.719] | 0.538 [0.520, 0.543] | 0.753 [0.746, 0.764] |
| **Infiltration** | 0.628 [0.539, 0.733] | 0.574 [0.450, 0.675] | 0.730 [0.629, 0.818] | 0.733 [0.632, 0.836] | **0.832 [0.708, 0.936]** | 0.629 [0.520, 0.759] | 0.663 [0.576, 0.759] | 0.580 [0.459, 0.700] |
| **Mass** | 0.602 [0.593, 0.610] | 0.578 [0.566, 0.587] | 0.623 [0.613, 0.631] | **0.626 [0.615, 0.637]** | 0.588 [0.578, 0.595] | 0.591 [0.580, 0.600] | 0.561 [0.546, 0.564] | 0.568 [0.557, 0.579] |
| **Nodule** | 0.687 [0.672, 0.703] | 0.626 [0.607, 0.643] | 0.781 [0.766, 0.795] | **0.826 [0.804, 0.840]** | 0.752 [0.734, 0.769] | 0.761 [0.746, 0.776] | 0.542 [0.521, 0.563] | 0.581 [0.564, 0.601] |
| **Pneumonia** | 0.617 [0.602, 0.636] | 0.518 [0.502, 0.532] | 0.680 [0.666, 0.700] | **0.699 [0.683, 0.718]** | 0.626 [0.611, 0.641] | 0.610 [0.596, 0.627] | 0.506 [0.496, 0.527] | 0.573 [0.562, 0.587] |
| **Pneumothorax** | 0.740 [0.730, 0.752] | 0.595 [0.581, 0.609] | 0.791 [0.783, 0.801] | **0.800 [0.791, 0.809]** | 0.700 [0.688, 0.710] | 0.695 [0.685, 0.709] | 0.516 [0.507, 0.533] | 0.663 [0.648, 0.675] |
| **Consolidation** | 0.782 [0.771, 0.797] | 0.691 [0.678, 0.705] | 0.856 [0.844, 0.868] | **0.872 [0.861, 0.884]** | 0.855 [0.844, 0.869] | 0.857 [0.842, 0.870] | 0.638 [0.628, 0.663] | 0.731 [0.717, 0.747] |
| **Edema** | 0.722 [0.707, 0.736] | 0.563 [0.543, 0.586] | **0.786 [0.772, 0.804]** | 0.782 [0.762, 0.800] | 0.708 [0.691, 0.728] | 0.672 [0.650, 0.694] | 0.580 [0.562, 0.615] | 0.587 [0.569, 0.610] |
| **Emphysema** | 0.690 [0.661, 0.713] | 0.540 [0.500, 0.577] | **0.747 [0.712, 0.791]** | 0.741 [0.693, 0.780] | 0.675 [0.635, 0.710] | 0.641 [0.603, 0.682] | 0.574 [0.541, 0.618] | 0.586 [0.552, 0.618] |
| **Fibrosis** | 0.628 [0.585, 0.675] | 0.6470 [0.606, 0.690] | **0.741 [0.704, 0.780]** | 0.718 [0.680, 0.766] | 0.680 [0.637, 0.723] | 0.692 [0.644, 0.749] | 0.613 [0.590, 0.667] | 0.628 [0.590, 0.686] |
| **Pleural Thickening** | 0.803 [0.784, 0.823] | 0.819 [0.801, 0.839] | 0.907 [0.892, 0.918] | **0.908 [0.894, 0.922]** | 0.876 [0.860, 0.893] | 0.863 [0.846, 0.878] | 0.753 [0.724, 0.775] | 0.751 [0.728, 0.776] |
| **Hernia** | 0.748 [0.734, 0.771] | 0.688 [0.670, 0.704] | 0.791 [0.777, 0.807] | **0.801 [0.787, 0.817]** | 0.737 [0.720, 0.753] | 0.751 [0.736, 0.772] | 0.658 [0.644, 0.687] | 0.717 [0.699, 0.737] |
| **Overall AUC** | 0.706 [0.683, 0.731] | 0.614 [0.586, 0.641] | 0.788 [0.765, 0.810] | **0.797 [0.772, 0.822]** | 0.743 [0.717, 0.768] | 0.723 [0.697, 0.752] | 0.587 [0.563, 0.618] | 0.640 [0.613, 0.670] |

**Table 13. NIH ChestX-Ray14 Results -** AUC across all diagnoses for Finetuning and Linear Probing.

| FINETUNING | ImageNet-Pretrained Baseline | MedAug | CXR-RePaiR-CLIP | REFERS | ConVIRT | GLoRIA | S2MTS2 | MoCo-CXR |
|---|---|---|---|---|---|---|---|---|
| Aortic enlargement | 0.856 [0.834, 0.876] | 0.790 [0.773, 0.810] | 0.850 [0.831, 0.869] | **0.898 [0.888, 0.913]** | 0.853 [0.830, 0.877] | 0.837 [0.817, 0.858] | 0.816 [0.799, 0.848] | 0.803 [0.779, 0.829] |
| Atelectasis | 0.795 [0.766, 0.838] | 0.775 [0.731, 0.826] | 0.814 [0.774, 0.857] | **0.844 [0.816, 0.881]** | 0.830 [0.802, 0.863] | 0.745 [0.684, 0.783] | 0.766 [0.715, 0.810] | 0.772 [0.714, 0.812] |
| Calcification | 0.786 [0.756, 0.815] | 0.809 [0.786, 0.831] | 0.810 [0.782, 0.835] | **0.842 [0.819, 0.856]** | 0.814 [0.781, 0.839] | 0.800 [0.768, 0.828] | 0.796 [0.747, 0.821] | 0.758 [0.724, 0.796] |
| Cardiomegaly | 0.893 [0.878, 0.909] | 0.829 [0.807, 0.853] | 0.894 [0.876, 0.911] | **0.935 [0.919, 0.947]** | 0.907 [0.885, 0.925] | 0.897 [0.877, 0.912] | 0.871 [0.846, 0.892] | 0.850 [0.828, 0.872] |
| Consolidation | 0.803 [0.758, 0.849] | 0.788 [0.740, 0.844] | 0.882 [0.858, 0.914] | **0.912 [0.877, 0.939]** | 0.848 [0.820, 0.891] | 0.815 [0.780, 0.858] | 0.778 [0.729, 0.834] | 0.795 [0.746, 0.847] |
| ILD | 0.804 [0.772, 0.847] | 0.805 [0.789, 0.829] | 0.840 [0.818, 0.872] | **0.853 [0.828, 0.879]** | 0.822 [0.797, 0.857] | 0.799 [0.775, 0.831] | 0.793 [0.769, 0.830] | 0.793 [0.768, 0.817] |
| Infiltration | 0.817 [0.768, 0.867] | 0.799 [0.730, 0.842] | 0.838 [0.788, 0.878] | 0.886 [0.829, 0.931] | **0.899 [0.839, 0.936]** | 0.868 [0.813, 0.909] | 0.852 [0.808, 0.899] | 0.874 [0.821, 0.912] |
| Lung Opacity | 0.792 [0.729, 0.834] | 0.768 [0.735, 0.801] | 0.825 [0.779, 0.855] | 0.823 [0.780, 0.865] | **0.829 [0.795, 0.867]** | 0.812 [0.778, 0.845] | 0.768 [0.722, 0.798] | 0.754 [0.712, 0.803] |
| Nodule/Mass | 0.747 [0.702, 0.784] | 0.756 [0.725, 0.790] | 0.800 [0.765, 0.838] | **0.858 [0.823, 0.886]** | 0.802 [0.771, 0.843] | 0.782 [0.752, 0.815] | 0.723 [0.687, 0.763] | 0.752 [0.718, 0.786] |
| Pleural effusion | 0.880 [0.841, 0.908] | 0.810 [0.782, 0.841] | 0.884 [0.847, 0.912] | **0.952 [0.923, 0.968]** | 0.942 [0.916, 0.960] | 0.948 [0.927, 0.959] | 0.866 [0.825, 0.893] | 0.824 [0.794, 0.848] |
| Pleural thickening | 0.821 [0.780, 0.845] | 0.808 [0.773, 0.836] | 0.829 [0.797, 0.855] | 0.862 [0.831, 0.893] | **0.868 [0.847, 0.890]** | 0.856 [0.829, 0.873] | 0.812 [0.767, 0.840] | 0.820 [0.789, 0.846] |
| Pulmonary fibrosis | 0.799 [0.767, 0.826] | 0.768 [0.745, 0.791] | 0.786 [0.755, 0.815] | 0.852 [0.830, 0.877] | **0.852 [0.835, 0.867]** | 0.835 [0.814, 0.856] | 0.777 [0.750, 0.800] | 0.786 [0.768, 0.815] |
| Other lesion | 0.821 [0.789, 0.852] | 0.786 [0.745, 0.817] | 0.832 [0.807, 0.857] | **0.833 [0.792, 0.857]** | 0.820 [0.791, 0.845] | 0.814 [0.787, 0.842] | 0.768 [0.729, 0.802] | 0.783 [0.754, 0.810] |
| Lung tumor | 0.743 [0.696, 0.803] | 0.745 [0.701, 0.793] | 0.828 [0.797, 0.858] | **0.881 [0.832, 0.918]** | 0.807 [0.774, 0.855] | 0.753 [0.708, 0.795] | 0.710 [0.657, 0.757] | 0.741 [0.689, 0.791] |
| Pneumonia | 0.818 [0.793, 0.841] | 0.794 [0.764, 0.821] | 0.876 [0.853, 0.895] | **0.913 [0.891, 0.929]** | 0.885 [0.868, 0.902] | 0.865 [0.847, 0.883] | 0.815 [0.792, 0.854] | 0.803 [0.773, 0.828] |
| Tuberculosis | 0.801 [0.760, 0.839] | 0.786 [0.754, 0.821] | 0.832 [0.797, 0.867] | 0.896 [0.862, 0.923] | **0.905 [0.882, 0.922]** | 0.865 [0.835, 0.895] | 0.812 [0.768, 0.846] | 0.814 [0.781, 0.844] |
| Other diseases | 0.827 [0.811, 0.849] | 0.829 [0.813, 0.846] | 0.831 [0.814, 0.847] | **0.874 [0.858, 0.889]** | 0.864 [0.850, 0.883] | 0.864 [0.855, 0.883] | 0.815 [0.797, 0.838] | 0.832 [0.819, 0.851] |
| No finding | 0.854 [0.844, 0.871] | 0.856 [0.843, 0.874] | 0.868 [0.854, 0.881] | **0.916 [0.907, 0.925]** | 0.909 [0.898, 0.919] | 0.899 [0.888, 0.910] | 0.849 [0.830, 0.865] | 0.860 [0.847, 0.874] |
| Overall AUC | 0.814 [0.780, 0.847] | 0.794 [0.763, 0.826] | 0.840 [0.811, 0.868] | **0.879 [0.850, 0.904]** | 0.859 [0.832, 0.886] | 0.836 [0.807, 0.863] | 0.799 [0.763, 0.833] | 0.801 [0.768, 0.832] |

| LINEAR PROBING | ImageNet-Pretrained Baseline | MedAug | CXR-RePaiR-CLIP | REFERS | ConVIRT | GLoRIA | S2MTS2 | MoCo-CXR |
|---|---|---|---|---|---|---|---|---|
| Aortic enlargement | 0.856 [0.834, 0.876] | 0.636 [0.596, 0.668] | 0.867 [0.846, 0.889] | **0.907 [0.892, 0.921]** | 0.780 [0.759, 0.808] | 0.673 [0.643, 0.705] | 0.646 [0.617, 0.687] | 0.606 [0.569, 0.645] |
| Atelectasis | 0.795 [0.766, 0.838] | 0.620 [0.555, 0.683] | 0.850 [0.811, 0.884] | **0.855 [0.812, 0.888]** | 0.740 [0.686, 0.805] | 0.682 [0.627, 0.739] | 0.575 [0.505, 0.643] | 0.490 [0.406, 0.565] |
| Calcification | 0.786 [0.756, 0.815] | 0.634 [0.598, 0.676] | **0.799 [0.774, 0.828]** | 0.791 [0.768, 0.820] | 0.660 [0.632, 0.691] | 0.605 [0.571, 0.649] | 0.624 [0.586, 0.672] | 0.580 [0.545, 0.631] |
| Cardiomegaly | 0.893 [0.878, 0.909] | 0.653 [0.621, 0.689] | 0.908 [0.890, 0.923] | **0.948 [0.938, 0.957]** | 0.870 [0.853, 0.887] | 0.779 [0.749, 0.804] | 0.720 [0.689, 0.751] | 0.712 [0.682, 0.740] |
| Consolidation | 0.803 [0.758, 0.849] | 0.722 [0.654, 0.780] | 0.921 [0.895, 0.944] | **0.931 [0.911, 0.953]** | 0.897 [0.863, 0.930] | 0.751 [0.702, 0.795] | 0.730 [0.683, 0.791] | 0.664 [0.608, 0.716] |
| ILD | 0.804 [0.772, 0.847] | 0.552 [0.515, 0.591] | **0.857 [0.834, 0.891]** | 0.818 [0.785, 0.868] | 0.752 [0.722, 0.791] | 0.661 [0.628, 0.706] | 0.698 [0.660, 0.736] | 0.617 [0.568, 0.663] |
| Infiltration | 0.817 [0.768, 0.867] | 0.653 [0.586, 0.731] | 0.860 [0.814, 0.909] | **0.885 [0.841, 0.924]** | 0.788 [0.722, 0.858] | 0.751 [0.690, 0.812] | 0.697 [0.609, 0.797] | 0.704 [0.619, 0.805] |
| Lung Opacity | 0.792 [0.729, 0.834] | 0.646 [0.593, 0.686] | 0.827 [0.786, 0.864] | **0.845 [0.810, 0.874]** | 0.744 [0.693, 0.792] | 0.633 [0.580, 0.678] | 0.647 [0.575, 0.710] | 0.562 [0.504, 0.615] |
| Nodule/Mass | 0.747 [0.702, 0.784] | 0.561 [0.522, 0.607] | 0.809 [0.774, 0.838] | **0.833 [0.798, 0.865]** | 0.686 [0.638, 0.731] | 0.601 [0.552, 0.650] | 0.533 [0.487, 0.583] | 0.544 [0.500, 0.604] |
| Pleural effusion | 0.880 [0.841, 0.908] | 0.758 [0.721, 0.799] | 0.940 [0.919, 0.958] | **0.955 [0.935, 0.974]** | 0.931 [0.902, 0.953] | 0.908 [0.883, 0.929] | 0.634 [0.559, 0.714] | 0.713 [0.651, 0.760] |
| Pleural thickening | 0.821 [0.780, 0.845] | 0.608 [0.561, 0.647] | **0.859 [0.828, 0.884]** | 0.854 [0.827, 0.883] | 0.720 [0.680, 0.765] | 0.653 [0.612, 0.693] | 0.589 [0.514, 0.638] | 0.626 [0.566, 0.675] |
| Pulmonary fibrosis | 0.799 [0.767, 0.826] | 0.554 [0.515, 0.609] | 0.820 [0.793, 0.848] | **0.847 [0.819, 0.874]** | 0.690 [0.654, 0.734] | 0.608 [0.576, 0.653] | 0.589 [0.542, 0.640] | 0.585 [0.534, 0.635] |
| Other lesion | 0.821 [0.789, 0.852] | 0.552 [0.498, 0.617] | 0.834 [0.796, 0.875] | **0.860 [0.833, 0.893]** | 0.706 [0.659, 0.749] | 0.615 [0.568, 0.679] | 0.601 [0.539, 0.661] | 0.563 [0.505, 0.634] |
| Lung tumor | 0.743 [0.696, 0.803] | 0.561 [0.511, 0.625] | 0.838 [0.793, 0.873] | **0.855 [0.806, 0.892]** | 0.745 [0.679, 0.818] | 0.653 [0.580, 0.709] | 0.583 [0.497, 0.643] | 0.513 [0.433, 0.574] |
| Pneumonia | 0.818 [0.793, 0.841] | 0.672 [0.638, 0.705] | 0.916 [0.900, 0.933] | **0.933 [0.914, 0.947]** | 0.867 [0.836, 0.892] | 0.765 [0.738, 0.790] | 0.702 [0.660, 0.742] | 0.644 [0.610, 0.689] |
| Tuberculosis | 0.801 [0.760, 0.839] | 0.577 [0.543, 0.631] | 0.848 [0.814, 0.885] | **0.885 [0.860, 0.919]** | 0.787 [0.757, 0.835] | 0.733 [0.695, 0.779] | 0.654 [0.611, 0.699] | 0.633 [0.589, 0.690] |
| Other diseases | 0.827 [0.811, 0.849] | 0.572 [0.550, 0.601] | 0.857 [0.843, 0.870] | **0.876 [0.861, 0.889]** | 0.642 [0.621, 0.669] | 0.565 [0.544, 0.594] | 0.610 [0.585, 0.640] | 0.555 [0.525, 0.583] |
| No finding | 0.854 [0.844, 0.871] | 0.543 [0.523, 0.571] | 0.903 [0.893, 0.916] | **0.923 [0.915, 0.934]** | 0.646 [0.627, 0.673] | 0.522 [0.500, 0.550] | 0.614 [0.592, 0.639] | 0.521 [0.494, 0.547] |
| Overall AUC | 0.814 [0.780, 0.847] | 0.615 [0.572, 0.662] | 0.862 [0.833, 0.890] | **0.878 [0.851, 0.904]** | 0.758 [0.721, 0.799] | 0.675 [0.635, 0.717] | 0.636 [0.584, 0.688] | 0.602 [0.550, 0.654] |

**Table 14. VinDr-CXR Results -** AUC across all diagnoses for Finetuning and Linear Probing.
